# Near, far, wherever you are: Phenotype-related variation in pharmacogenomic effect sizes across the psychiatric drug literature

**DOI:** 10.1101/2025.09.23.25336442

**Authors:** Siobhan K. Lock, Djenifer B. Kappel, Jude Hutton, Emily Simmonds, Sophie E. Legge, Michael C. O’Donovan, Antonio F. Pardiñas

## Abstract

**Background:** Pharmacogenomics is viewed as one route to minimising inter-individual variability in drug response. However, uptake in psychiatry is slower than in other medical fields, so assessing evidence for psychiatric genotype-drug pairs and understanding what influences the magnitude of these effects is essential.

**Methods:** We performed a systematic search for studies investigating pharmacogenomic variation in the context of antipsychotic and antidepressant use. Outcomes varied, including those related to drug bioavailability (“proximal”) or side effects, symptom severity, and other treatment outcomes (“distal”). We performed a meta-analysis moderated by outcome type to quantify the average pharmacogenomic effect size across proximal and distal outcomes and assess whether they differ from one another. We developed a companion dashboard that allows users to explore the dataset and perform simplified meta-analyses, power calculations, and Bayesian shrinkage analyses based on drugs, enzymes, and outcomes of interest.

**Results:** We analysed 2,092 standardised mean differences (SMDs) from 184 studies, finding evidence that pharmacogenomic effect sizes for proximal outcomes were significantly larger than distal (Δβ = −0.206 [95% CI −0.291 – −0.121], *p* = 5×10^−6^). This trend was consistent across sub-groups restricted to the most common gene-drug pairings in the dataset. Power calculations for hypothetical future studies using two-sample t-tests showed that, to attain at least 80% statistical power, analyses of distal outcomes require a larger sample size than proximal outcomes.

**Discussion:** We demonstrate that pharmacogenomic effect sizes are significantly larger for proximal outcomes related to pharmacokinetics than for distal outcomes related to efficacy and toxicity. Understanding how the biological mechanisms underlying different outcomes might impact pharmacogenomic effect sizes could help to inform participant recruitment for future psychiatric pharmacogenomic studies, alongside the development of pharmacogenomic guidelines for psychiatric medications.

**Funding:** This work was supported by a PhD studentship from Mental Health Research UK (SKL). Authors were also supported by grants from the Medical Research Council (SEL, MCOD, AFP; MR/Y004094/1 and MR/Z503745/1), the European Union’s Horizon 2020 programme (DBK, MCOD, AFP; #964874), the European Union’s Horizon 2021 programme (AFP, #101057454), and the Dementia Research Institute (ES; UKDRI supported by the Medical Research Council (UKDRI-3206), Alzheimer’s Research UK, and Alzheimer’s Society). The authors declare no conflict of interests.

## Introduction

The effects of most licensed therapeutic drugs vary even when prescribed to individuals for the same indication. Moreover, real-world effectiveness often falls below the expectations set by clinical trials, an “efficacy-effectiveness gap” that pervades medicine (Eichler et al., 2011). Pharmacological effects can lead to huge benefits for some individuals, be barely noticeable for others, and for a few even cause a net negative impact through adverse drug reactions (ADRs). Problems with the external validity of randomised controlled trials, diverse healthcare and drug use practices, or patient-level variation in drug response are all likely contributors to the gap (Groenwold, 2021). In psychiatry, where “treatment resistant” symptoms affect a large proportion of patients (Howes et al., 2022), these issues have been systematically raised and examined. While there is no evidence that common psychotropics or even psychotherapy interventions are less efficacious than drugs prescribed for non-psychiatric conditions (Leucht et al., 2012; Huhn et al., 2014), the development and refinement of psychiatric drugs is notoriously slow compared to other fields (Nutt, 2025), partly because many mechanisms of action involved in psychopharmacology and their precise effect on disease processes and neurobiology remain unclear (Huda, 2019; Howes et al., 2022).

Substantial efforts in psychiatric research are dedicated to more accurately predicting treatment response to current drugs, with aims of eventually lessening the efficacy-effectiveness gap (Dellen, 2024). One route to accomplishing this task is stratified medicine or “precision psychiatry”, which involves identifying patient subgroups with respect to their characteristics or response to drugs (Bell, 2014). While a myriad of factors (e.g., environmental, physiological, genomic) are known to affect treatment response in psychiatry (Stern et al., 2018), genetic variation is of particularly strong interest for developing stratified approaches as it is identifiable and stable from birth. Which genetic variants affect response to drugs, and whether they influence pharmacokinetic or pharmacodynamic processes, is the topic of study of pharmacogenomics (Pirmohamed and Park, 2001). Advances in this field have led to algorithms, developed by expert groups (Klein and Ritchie, 2018) or industry (Behera et al., 2025), that can inform screening for haplotypes known to influence the activity of drug metabolising enzymes (classically termed “star alleles”), and infer how carriers might respond to a given drug. While the research sustaining the psychiatric arm of this discipline has seen a large growth in activities in recent years, and an increasing number of prospective trials are being funded, most promising results are still confined to the scientific literature and have not yet been translated into clinical interventions (Bousman et al., 2023a).

Translational efforts need to be based on robust research evidence, and pharmacogenomics has a number of international expert consortia routinely assessing any pharmacogenomic reports linked to licensed drugs (Whirl-Carrillo et al., 2021). Indeed, for many drug-gene pairs, pharmacogenomic effects have been investigated broadly, from simple molecular processes to complex patient-reported outcomes. While any of these reports could be used by regulators in deciding whether to recommend an implementation of pharmacogenomic testing (Wu, 2015), genetic evidence related to pharmacodynamic phenotypes such as drug response and ADRs tends to be considered the most likely to lead to a clinically useful and cost-effective intervention (Hughes, 2018).

This perception is worth emphasising. In psychiatry, robust associations of pharmacogenomic variants with pharmacodynamic outcomes are rare, and pharmacogenomic guidelines for psychotropics are mostly based on studies of drug metabolism and other indicators of drug pharmacokinetics (Bousman et al., 2020). A potential explanation for this asymmetry is that, outside of severe ADRs, pharmacodynamic phenotypes seem akin to complex traits. They are multifactorial, polygenic, and require large samples for accurately estimating genetic effects, given these are often weak (Roden et al., 2019). In contrast, pharmacokinetic phenotypes seem to fit oligogenic inheritances with moderate-to-high heritabilities, and are therefore more tractable for genetic discovery studies (Roden et al., 2006; Ingelman-Sundberg and Molden, 2025). This echoes classic discussions in psychiatry about the use of “endophenotypes” : quantitative measures of biological processes relevant to psychiatric disorders that show stronger associations with genetic variants than the disorders themselves (Gottesman and Gould, 2003). Endophenotypes are meant to be surrogates of some of the elements that characterise multifactorial disorders, in the same way that pharmacokinetic processes are important drivers of pharmacodynamics, but not the only ones (Simon and von Fabeck, 2025). This implies the expectation that the effects of any given genetic variant are largest on phenotypes that closely reflect the biological processes that it directly causes or moderates. On the other hand, effect sizes become diluted as phenotypes move further away from basic molecular mechanisms and become influenced by other factors, genetic and otherwise.

In this paper, we aim to quantify effects of pharmacogenomic variation through this framework, initially postulated in research on metabolic control (Kacser and Burns, 1981). Borrowing from the “proximal-distal continuum” concept of health outcomes research (Brenner et al. 1995), we term “proximal outcomes” as those based on quantitative biological measures, which tend to be mechanistically closer to genetic variation. On the other end, “distal outcomes” are those usually captured by clinical phenotyping, being often further away from any single genetic effect and subject to multifactorial influences. We implemented this classification on a series of meta-analyses of pharmacogenomics reports focused on enzymes important to the metabolism of antidepressant and antipsychotic drugs (i.e., the cytochrome P450, or CYP, family of enzymes). While this literature is necessarily heterogeneous, investigating many drugs, enzymes, and outcomes, our aim was to curate a corpus of data that can be used for diverse applications, including the design and interpretation of psychiatric pharmacogenomic studies. If indeed genetic effects differ between proximal and distal outcomes, identifying even a wide range of plausible effect sizes for each category could assist with accurately estimating the statistical power of future basic and translational research (Huang et al., 2020), as well as flagging potential errors or biases in existing studies (Gelman and Carlin, 2014).

## Methods

### Literature Search

We searched PUBMED, Cochrane, and SCOPUS databases for literature published between 1996 (when standardised pharmacogenomic nomenclature was introduced) and November 2024. Briefly, the search looked across titles for the following key terms in the format [pharmacogenomic term] AND [drug term] AND [outcome term] AND NOT [‘review’ / ‘guidelines’]. The full list of search terms is found in the **Supplementary Materials**. Records returned from the three databases were combined, and duplicate entries were removed (**Figure 1**).

**Figure 1.**
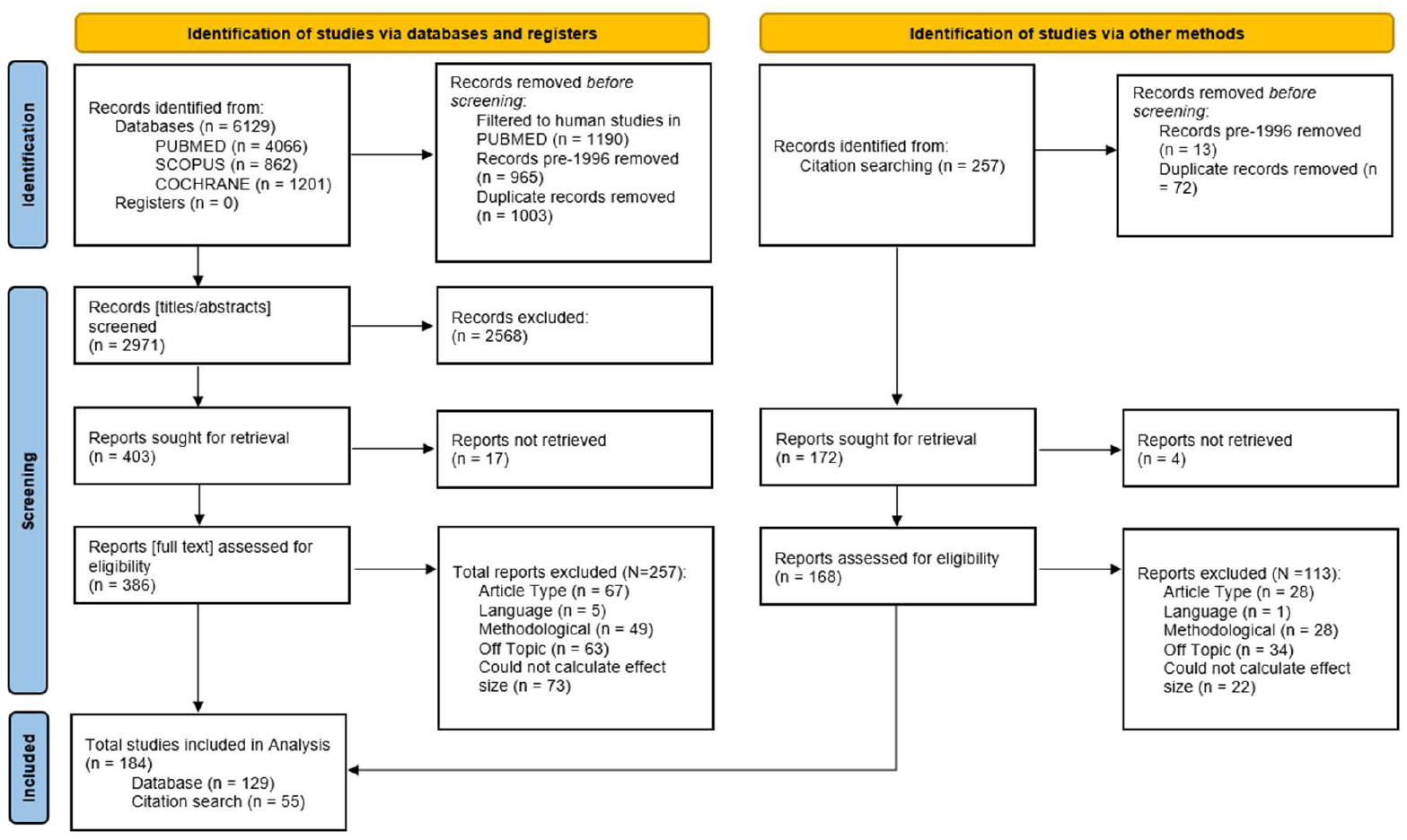
Flowchart showing the number of records identified from the literature search, the numbers following screening, retrieval, and finally, the number of records included in the final analysis. Flowchart template was adapted from the Preferred Reporting Items for Systematic reviews and Meta-Analyses (PRISMA; Moher et al. 2010).

### Study Selection

We compiled studies investigating associations of genotype-based pharmacogenomic variables with a range of outcomes in individuals taking psychiatric drugs. Broadly, these outcomes included measures of drug or metabolite bioavailability or clearance, drug doses, and measures of clinical response or ADRs (**Table 1**). We focussed on pharmacogenomic variation (i.e., genetics-inferred metabolism phenotypes, pharmacogenomic star alleles) in the CYP family of proteins given their role in the metabolism of many widely prescribed drugs (Zhao et al., 2021), including most psychotropics (Spina and de Leon, 2015).

**Table 1.** Examples of phenotypes included in the meta-analyses and whether they were rated as proximal or distal outcomes.

| Proximal Outcomes | Distal Outcomes |
| --- | --- |
| Active moiety pharmacokinetics (e.g., concentration, half-life, clearance, AUC) | Adverse Drug Reactions |
| Drug pharmacokinetics (e.g., concentration, half-life, clearance, AUC) | Drug dose |
| Metabolite pharmacokinetics (e.g., concentration, half-life, clearance, AUC) | Treatment outcomes (e.g., remission, hospitalisation, treatment discontinuation, medication switching) |
| Metabolic ratios | Symptom severity |

Titles and abstracts were screened first to determine eligibility based on fit to the research topic of this study. For reports passing this screening, we retrieved the full text and supplementary material if available. After full text inspection, we included reports in our analyses that were (i) original research articles, (ii) in English language, or able to be translated without ambiguities by the authors, (iii) were methodologically relevant, (iv) investigated an outcome, enzyme, or drug of interest, and (v) contained sufficient information to extract or infer effect sizes (see “Statistical Analysis”). We excluded studies conducted *in vitro*, using population modelling, or investigating drug-drug interactions. Studies in which pharmacogenomic information was not used (i.e., phenotyping by probe drug) or those investigating combinatorial metabolism phenotypes were also excluded. Where effect size information was shown graphically but was not explicitly mentioned in the report text, it was extracted from plots using a free online application (PlotDigitizer: Version 3.1.6, 2025). Finally, we also examined the reference lists of included studies for any relevant work that might have been missed by the initial literature search and screened these references as above (**Figure 1**).

### Statistical Analysis

Data were analysed in R v4.4.0 in R Studio 2024.04.0 Build 735 (R Core Team, 2021). Based on the information available in the eligible reports, we derived standardised mean differences (SMDs). In the context of this study, SMDs refer to the difference in effect between a group of individuals with an atypical CYP metabolism phenotype versus a normal metabolism phenotype. We also extracted SMDs for the effects of individual pharmacogenomic variants against wild-type alleles when these were reported. To calculate SMDs, group means and standard deviations were the preferred inputs. Where median and (interquartile) ranges were reported, approximate means and standard deviations were calculated using methods outlined by Wan et al. (2014). These were converted to SMDs using the *esc* ‘effect size conversion’ R package (Lüdecke, 2018). Other data reported (e.g., number of observations, Odds Ratios, and *t* statistics, in order of preference) were converted using the *esc* package. Full information about these calculations is provided in the **Supplementary Materials**.

Additional details that were extracted from each report included: authors, year of publication, drug, enzyme, assessed outcome, and pharmacogenomic information. Where possible, pharmacogenomic information was extracted as metabolism phenotypes, or alternatively converted to them using on PharmGKB and CPIC reference tables (**Supplementary Materials**). When conversion was not possible, information was left in the same format as reported. For classifying outcomes, a binary variable was created, as described in **Table 1**. In summary, pharmacokinetic outcomes (e.g., relating to bioavailability or clearance of a drug or metabolite) were rated as “proximal” . Outcomes were alternatively labelled “distal” if they related to pharmacodynamics or potential consequences of altered metabolism; these included, for example, reports of drug doses, experiences of adverse drug reactions (ADRs), or ratings of symptom severity.

Our primary analysis utilised the full set of calculated effect sizes. To account for heterogeneity between drugs, enzymes, and pharmacogenomic measures analysed, we used absolute effect sizes as the outcome variable, and the outcome binary rating (i.e., proximal/distal) as the moderator variable. We included a random intercept for effect size ID, and study ID, and a random slope for the moderator, with an unstructured variance-covariance matrix. As a sensitivity analysis, we repeated the primary meta-analysis on gene-drug subgroups to determine whether findings relating to proximal and distal effect sizes were replicable across subsets of the data. Gene-drug pairings with at least one hundred effect sizes were selected, including CYP2D6-risperidone (N = 412), CYP2C19-escitalopram (N = 215), CYP2D6-aripiprazole (N = 120), and CYP2D6-haloperidol (N = 106). For these analyses, we used a simplified random effects structure of effect size ID nested within study ID.

As a secondary analysis, we tested for whether the outcome phenotype (“proximal” or “distal”) influenced the effects of genetics-inferred enzyme activity across distinct metabolism phenotypes. This analysis included only effect sizes for which pharmacogenomic information was available as, or could be converted to, metabolism phenotypes. We extended the primary meta-analysis model to include an interaction term between rating and metabolism phenotype as the moderators, keeping the same random-effects structure. Comparisons between estimates of metaboliser phenotype effect sizes were performed using Wald-type tests. Holm’s method was used to correct for multiple comparisons.

We used *metafor* (Viechtbauer, 2010) to fit multi-level, multivariate meta-analyses. In all analyses, we used a three-level correlated and hierarchical effects model. To account for dependency in our data, we imputed a variance-covariance matrix in *metafor* with positive correlations between clusters of effects (ρ = 0.6). We also performed robust variance estimation based on the *clubSandwich* package (Pustejovsky, 2024) to further account for this dependency via a sandwich estimator with bias reduced-linearisation for small-sample correction. We tested models using different random effect structures, alongside different parameters of ρ to assess how these affected our analysis, also using profile likelihood plots to assess parameter identifiability and potential model over-parametrisation (see **Supplementary Materials**). We fit two meta-analyses, one without the intercept to attain effect estimates for both proximal and distal outcomes, and one with an intercept to test whether proximal and distal effect estimates are significantly different. Finally, due to concerns that assessing traditional funnel plots using the SMD can lead to false positives when checking for publication bias, we additionally plot the SMD against 1/√N (Zwetsloot et al., 2017). We formally tested for publication bias using an adjusted version of Egger’s Test that accounts for the multi-level nature of the data, also using 1/√N in place of the standard error moderator (Rodgers and Pustejovsky, 2021).

### Applications

We conducted power analyses using *pwr* (Champely, 2020) to assess how sample sizes required to attain 80% power in pharmacogenomics research vary depending on the phenotype assessed. Effect sizes used were the proximal and distal effect estimates obtained from the primary meta-analysis and their robust 95% confidence intervals.

We also used the SMDs generated as part of this meta-analysis to model the potential inflation across effect sizes for proximal and distal outcomes (i.e., winner’s curse; Zöllner and Pritchard 2007). As described in the **Supplementary Material**, results from our models can be used as prior distributions to generate posterior bias-reduced estimates of pharmacogenomic effects in existing and future studies.

Finally, we developed an interactive dashboard allowing users to filter, browse, visualise, and download data collected for this meta-analysis. This allows interested readers to access the effect size data in a user-friendly manner, alongside acting as a resource to help researchers to plan their future studies into pharmacogenomic variation and psychiatric medicine through a basic power calculation. The dashboard also allows users to fit three-level correlated and hierarchical effects meta-analyses on the filtered data to allow for an initial assessment of the pooled effect sizes for genes, drugs, and enzymes of interest. The dashboard was built using the R *shiny* package (Chang et al., 2025).

## Results

This meta-analysis was performed in line with the Preferred Reporting Items for Systematic reviews and Meta-Analyses (PRISMA; Moher et al. 2010) guidelines (**Supplemental Materials**). **Figure 1** describes how the final list of studies was ascertained. Briefly, we identified and screened for eligibility 6,129 entries from the literature search, of which 386 full texts were selected to be assessed for inclusion. Following this, we identified an additional 257 records from citation searching, of which 168 were selected to have full texts assessed for inclusion. In total, we calculated 2,119 effect sizes from 184 studies (N = 129 from the literature search; N = 55 from citation searching). After excluding extreme values (SMD >= 5 or SMD <= −5), 2,092 effect sizes remained.

Over half (63%) of analyses were performed on samples recruited in European countries, with the next largest group (23%) coming from Asian countries. The most frequently studied drugs were risperidone (20%) and escitalopram (12%). The most studied enzymes were CYP2D6 (57%) and CYP2C19 (30%). There were similar numbers of proximal (56%) and distal (44%) outcomes across the analyses. A full description of the dataset is found in **Table 2**, below.

**Table 2.** Descriptive information about the analyses included in the effect size data. For each variable, number of studies and effect sizes are reported. The mean of the Standardised Mean Difference (SMD) and absolute Standardised Mean Difference (abs(SMD)) for each group are also given.

| Variable | N (effect sizes) (%) | N (studies) (%) | Mean SMD | Mean abs(SMD) |
| --- | --- | --- | --- | --- |
| Outcome |  |  |  |  |
| Proximal | 1,162 (56%) | 124 (53.68) | 0.197 | 0.702 |
| Distal | 930 (44%) | 107 (46.32) | 0.062 | 0.393 |
| Continent |  |  |  |  |
| Asia | 475 (23%) | 69 (37.5) | 0.236 | 0.641 |
| Europe | 1,318 (63%) | 91 (49.46) | 0.127 | 0.583 |
| Mixed | 4 (0.2%) | 1 (0.54) | 0.083 | 0.083 |
| North America | 213 (10%) | 18 (9.78) | 0.04 | 0.453 |
| Oceania | 72 (3.4%) | 3 (1.63) | 0.006 | 0.096 |
| South America | 10 (0.5%) | 2 (1.09) | -0.191 | 0.531 |
| Africa | 0 (0%) | 0 (0) | NA | NA |
| Drug Studied |  |  |  |  |
| Antidepressant | 829 (40%) | 60 (37.97) | 0.143 | 0.629 |
| Antipsychotic | 1,019 (49%) | 98 (62.03) | 0.132 | 0.573 |
| Unknown/Multiple | 244 (12%) | 34 (17.71) | 0.14 | 0.312 |
| Enzyme Studied |  |  |  |  |
| CYP1A2 | 139 (6.6%) | 19 (8.41) | -0.038 | 0.299 |
| CYP2B6 | 35 (1.7%) | 3 (1.33) | -0.046 | 0.428 |
| CYP2C19 | 621 (30%) | 45 (19.91) | 0.044 | 0.413 |
| CYP2C9 | 46 (2.2%) | 6 (2.65) | 0.101 | 0.359 |
| CYP2D6 | 1,184 (57%) | 145 (64.16) | 0.234 | 0.693 |
| CYP3A4/5 | 67 (3.2%) | 8 (3.54) | -0.226 | 0.464 |
| Metabolism Phenotype |  |  |  |  |
| Intermediate | 807 (48%) | 130 (39.63) | 0.246 | 0.583 |
| Poor | 425 (25%) | 68 (20.73) | 0.313 | 0.834 |
| Rapid | 113 (6.8%) | 18 (5.49) | -0.118 | 0.236 |
| Ultrarapid | 327 (20%) | 50 (15.24) | -0.129 | 0.456 |
| Unknown | 420 | 62 (18.9) | 0.025 | 0.429 |

### Meta-Analysis of Proximal and Distal Outcomes

As shown in **Figure 2**, we found evidence for a significant effect of pharmacogenomic variation on proximal (β = 0.486 [95% CI 0.415 – 0.557], *p* = 6×10^−25^) and distal outcomes (β = 0.28 [95% CI 0.219 – 0.34], *p* = 1×10^−13^). The intercept model showed that the proximal effect size was indeed significantly larger than the distal effect size (Δβ = −0.206 [95% CI −0.291 – −0.121], *p* = 5×10^−6^). Most of the variance in our dataset was at a within-study level (σ^2^ = 0.163). Between-study variance was larger for proximal outcomes (τ^2^ = 0.064), than for distal outcomes (τ^2^ = 0.017).

**Figure 2.**
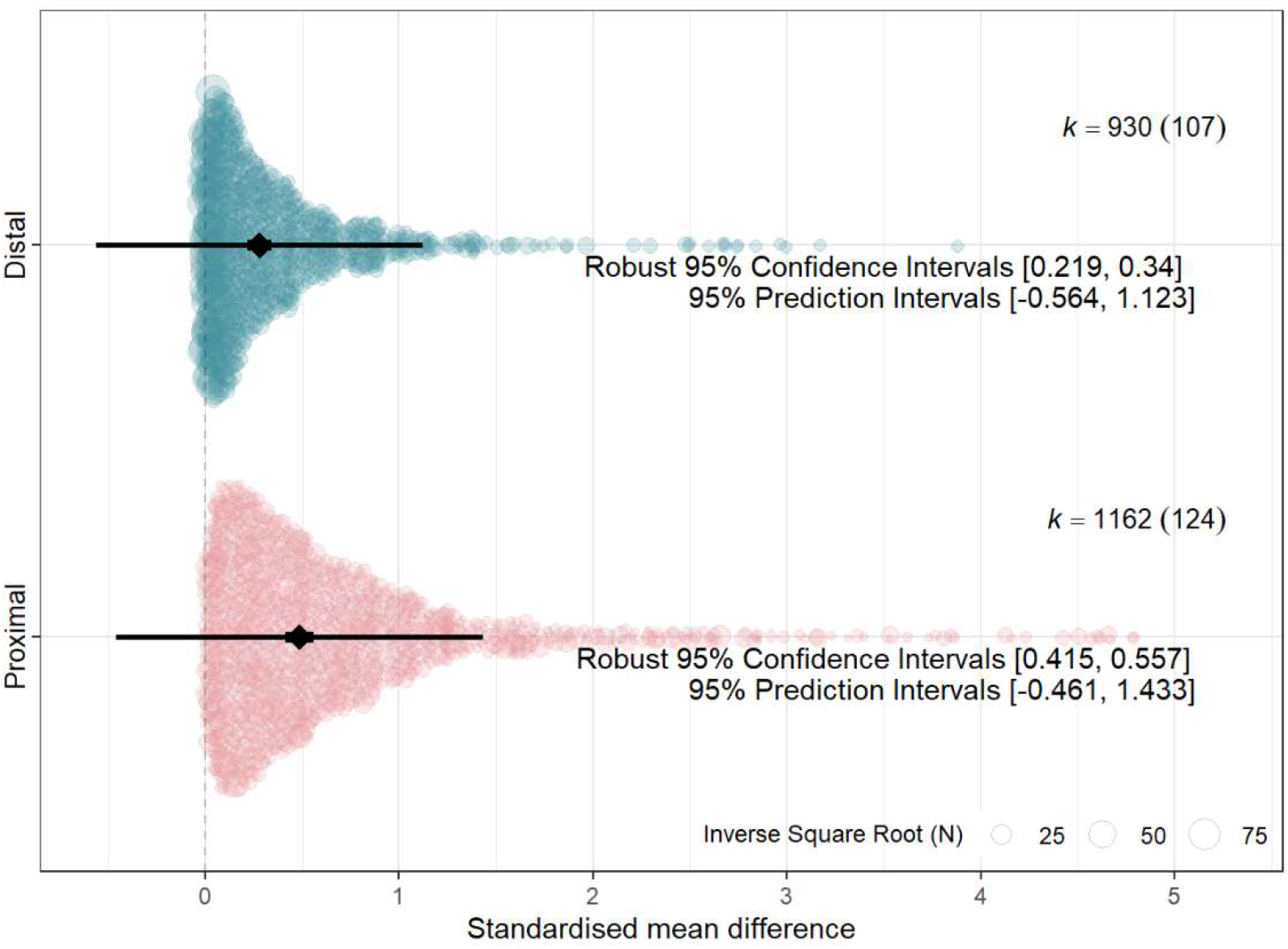
Orchard plot (Nakagawa et al., 2021) showing the distribution of effect sizes (Standardised Mean Difference, SMD) across proximal and distal outcomes. Size of points is determined by a sample-size based measure of precision (1/√N). Pooled estimates for proximal and distal effect sizes are represented by the black diamond, with error bars representing robust 95% confidence intervals (thick) and 95% prediction intervals (thin); see IntHout et al., (2016) for more information. Number of effect sizes is represented by *k*, with number of studies in brackets. Note that while an absolute scale was used for the effect sizes included in the meta-analysis, standard procedures were used for calculating prediction intervals, in which the lower bound is not limited at zero.

### Sensitivity Meta-Analyses: Across Gene-Drug Pairings

We repeated the primary analysis in subgroups restricted to the most common gene-drug pairings (i.e., those with at least 100 effect sizes available). Proximal effect sizes were significantly larger than distal effect sizes in the Risperidone-CYP2D6 group (Δβ = −0.206 [95% CI −0.361 – −0.052], *p* = 0.013), and the Aripiprazole-CYP2D6 group (Δβ = −0.301 [95% CI −0.591 – −0.011], p = 0.044). We did not observe a significant difference between the effect sizes in the haloperidol-CYP2D6 group (Δβ = −0.294 [95% CI −0.602 – 0.014], *p* = 0.057) and the escitalopram-CYP2C19 group (Δβ = −0.213 [95% CI − 0.716 – 0.29], *p* = 0.345). However, the directions of all effects were consistent with the primary analysis. Results for effect sizes for proximal and distal outcomes, separately, across each of the gene-drug subgroups are given in the **Supplementary Materials**.

### Secondary Analysis: Differences across Metabolism Phenotypes

We fit a meta-analysis restricted to effect sizes for which metabolism phenotype information was available. We tested for moderation effects via the addition of an outcome-metabolism phenotype interaction term to the model, comparing the effect of an atypical metabolism phenotype (i.e., Poor, Intermediate, Rapid, or Ultrarapid) versus the normal metabolism phenotype for either proximal, or distal outcomes. We included 1672 effect sizes from 146 different studies with results shown in **Figure 3** and the **Supplementary Materials**.

**Figure 3.**
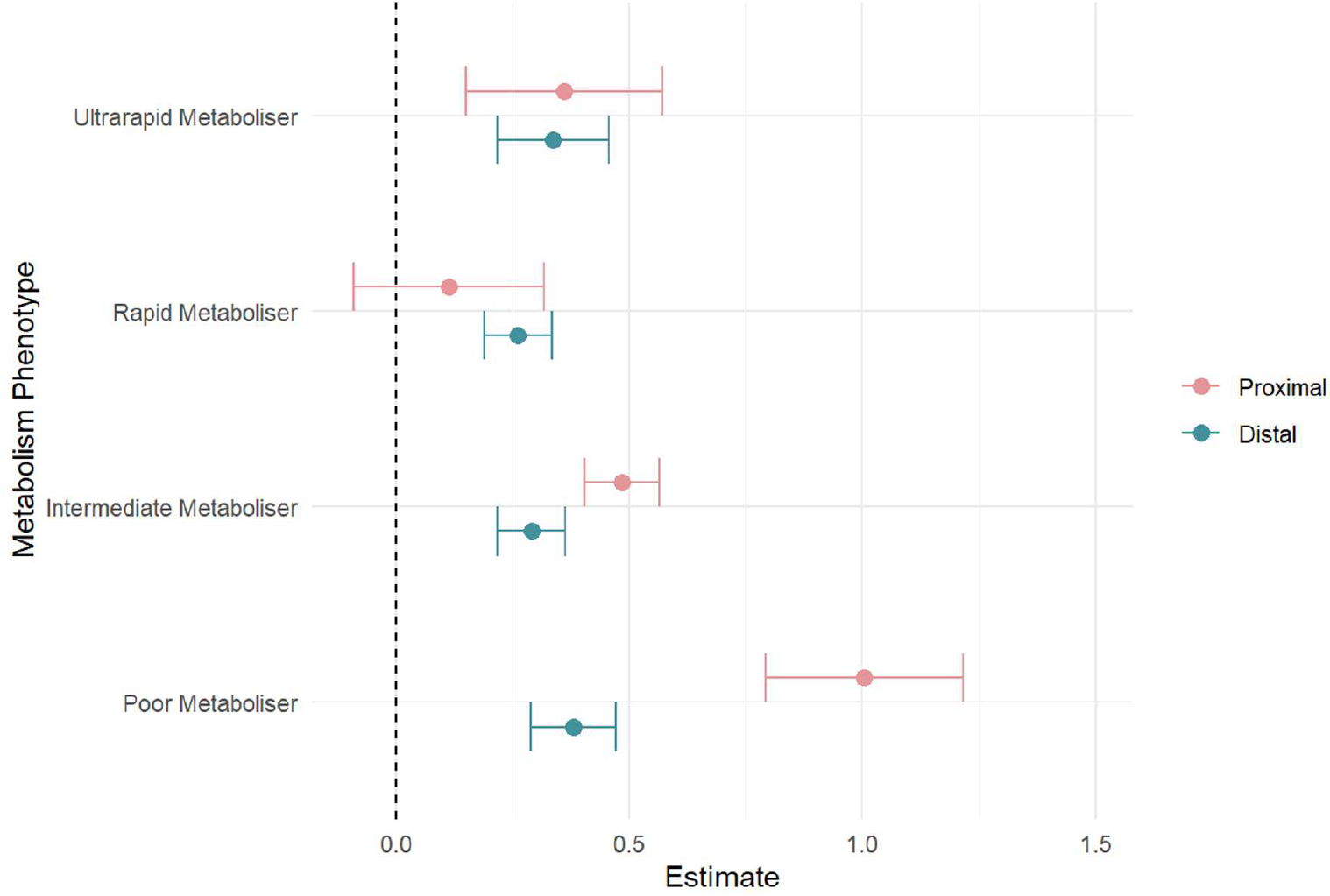
Forest plot showing absolute effect sizes of pharmacogenomic metabolism phenotypes on proximal and distal outcomes. Point shows the effect estimates for proximal (pink) and distal (blue) outcomes, error bars show robust 95% confidence intervals. Effect estimates reflect that of an atypical metabolism phenotype (i.e., Poor, Intermediate, Rapid, or Ultrarapid) versus the normal metabolism phenotype for either proximal, or distal outcomes.

We tested for differences between model estimates using Wald-type tests. Reported *p* values are corrected for multiple comparisons with Holm’s method. Effect sizes for proximal outcomes were significantly larger than distal outcomes for the poor metabolism phenotype (β = −0.623 (SE = 0.105), *p* = 2×10^−6^) and the intermediate metabolism phenotype (β = −0.194 (SE = 0.046), *p* = 2×10^−4^). There was no significant difference between proximal and distal estimates for both the rapid (β = 0.148 (SE = 0.094), *p* = 0.288) and ultrarapid (β = −0.023 (SE = 0.112), *p* = 0.836) metaboliser statuses. When assessing whether metabolism phenotypes had distinct effects within outcomes, we observed that the estimate for the poor metabolism phenotype was significantly different to all other phenotypes for proximal outcomes (**Table 3**). The effect size of an intermediate metabolism phenotype was also significantly larger than that of a rapid metabolism phenotype. For distal outcomes, there were no significant differences between the effect estimates of metabolism phenotypes (**Table 4**). As in the primary analysis, the largest variance component was at the within-study level (σ^2^ = 0.171), with greater between-study heterogeneity for proximal outcomes (τ^2^ = 0.077) than distal outcomes (τ^2^ = 0.024).

**Table 3.**
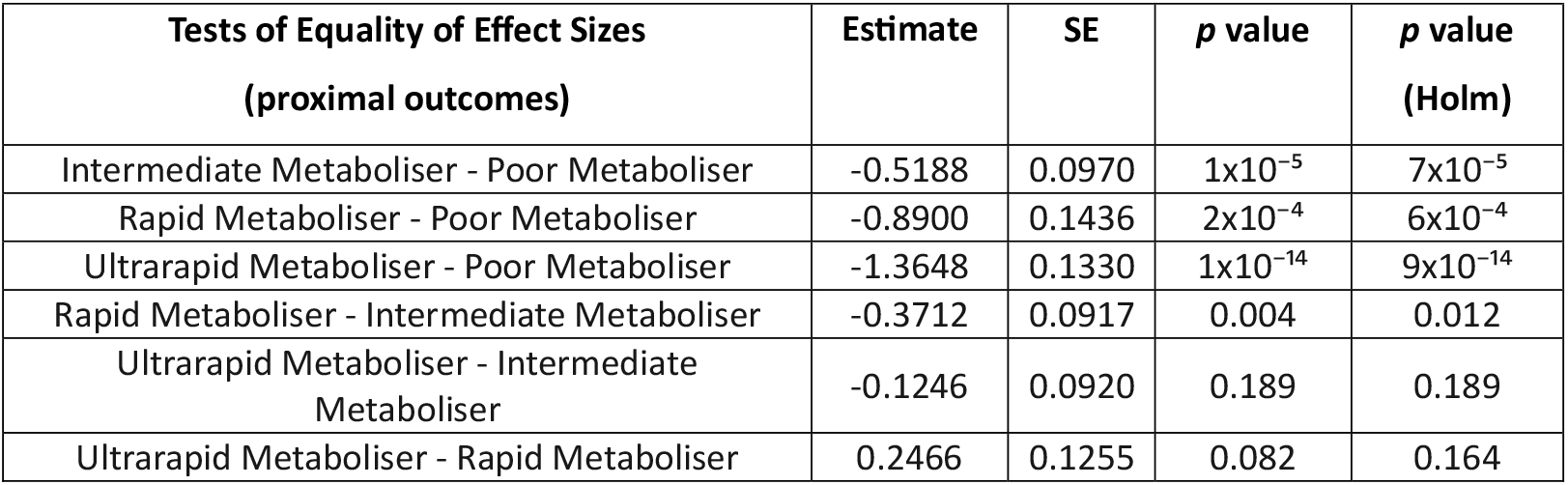
Results of ANOVA comparing whether proximal effect sizes for two different metabolism phenotypes (shown in the test column) are significantly different from each other.

**Table 4.** Results of ANOVA comparing whether distal effect sizes for two different metabolism phenotypes (shown in the test column) are significantly different from each other.

| Tests of Equality of Effect Sizes<br>(distal outcomes) | Estimate | SE | *p* value | *p* value<br>(Holm) |
| --- | --- | --- | --- | --- |
| Intermediate Metaboliser - Poor Metaboliser | -0.0894 | 0.0368 | 0.0250 | 0.1270 |
| Rapid Metaboliser - Poor Metaboliser | -0.1184 | 0.0353 | 0.0140 | 0.0830 |
| Ultrarapid Metaboliser - Poor Metaboliser | -0.0434 | 0.0414 | 0.3140 | 0.8940 |
| Rapid Metaboliser - Intermediate Metaboliser | -0.0290 | 0.0301 | 0.3750 | 0.8940 |
| Ultrarapid Metaboliser - Intermediate Metaboliser | 0.0460 | 0.0423 | 0.2980 | 0.8940 |
| Ultrarapid Metaboliser - Rapid Metaboliser | 0.0750 | 0.0425 | 0.1270 | 0.5100 |

### Applications

Based on the effect sizes generated in the primary meta-analysis, across all drugs and genes assessed, we generated curves to visualise the relationship between sample size and statistical power. These values are given as N per group for a two-sample *t*-test and are based on the average pharmacogenomic effect sizes estimated for proximal and distal outcomes (**Figure 4**). For a proximal outcome, the optimum sample size for 80% power was estimated at 68 individuals per group (range: 52 - 93), where each group constitutes a specific genotype or metabolism phenotype. For a distal outcome, the optimum sample size for 80% power was estimated 202 individuals per group (range: 137 - 328). Similarly, the Bayesian analysis supports a larger shrinkage factor (also called “exaggeration ratio”) across most of the z-score distribution for distal outcomes (mean = 1.97) than for proximal outcomes (mean = 1.28). Assuming the simplest scenario of equal unadjusted effect sizes, this suggests that posterior estimates for distal outcomes will be smaller than that for proximal outcomes in most cases (**Supplemental materials**). Finally, we developed a companion Shiny App allowing for the visualisation of effect size distributions and power curves. We also included the ability for users to perform their own exploratory meta-analyses using the full dataset, or filtered to whichever enzyme, drug, and outcome combinations desired. This also generates probability distributions based on the filtered data and a calculator for estimating posterior effect sizes that account for winner’s curse.

**Figure 4.**
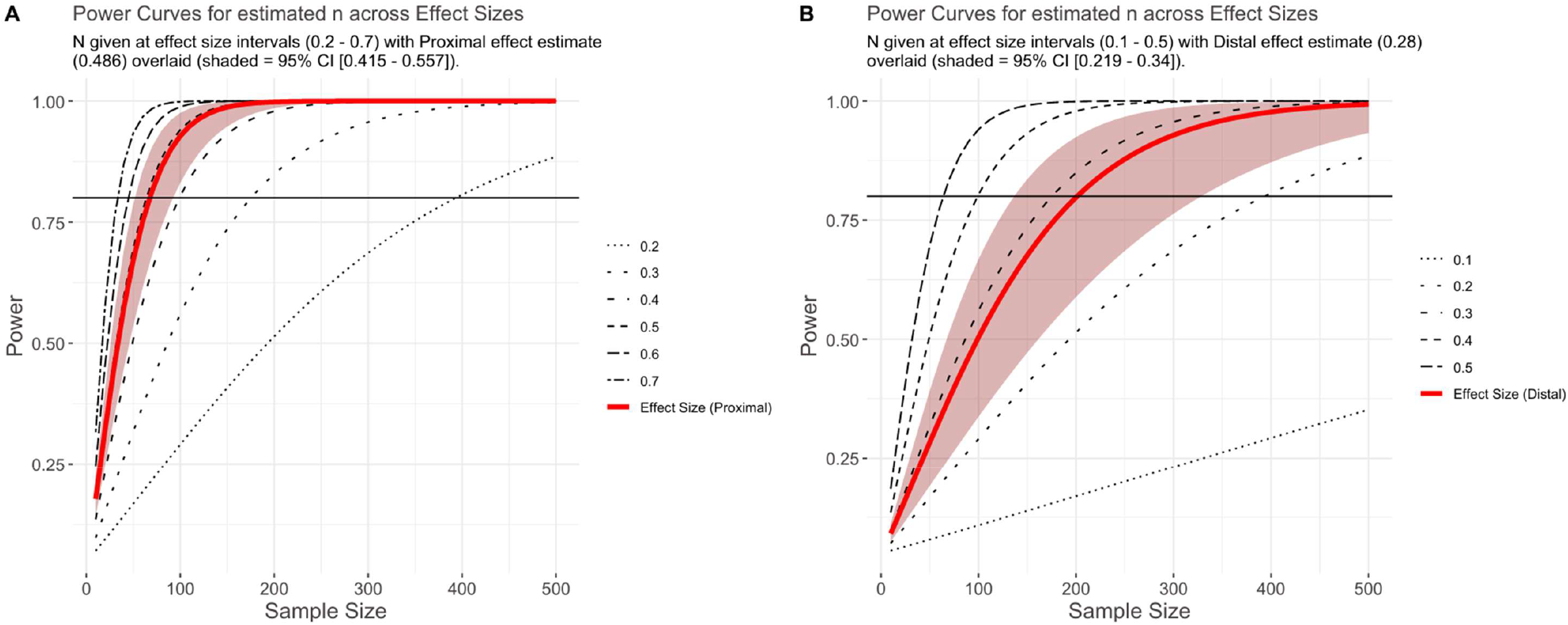
Plot shows power at a given effect size and sample size. Panels show power curves based on effect sizes generated for proximal (Panel A) and distal (Panel B) effect sizes (and robust 95% CI) from the primary meta-analysis. Black horizontal line indicates 80% power.

## Discussion

Here, we assess and quantify the effect size of pharmacogenomic variation across a large spectrum of phenotypes related to psychiatric drugs. We report on 2,092 effect sizes from 184 studies, demonstrating larger effects of pharmacogenomic variation on proximal outcomes than on distal outcomes. We showcase how the collected corpus of studies can be used to inform future pharmacogenomics research and study design and provide an interactive web app for browsing the raw data, facilitating the reproduction of our analyses or their adaptation to outcomes or drugs of specific interest for other researchers.

Our primary finding was that, as expected, the effect for pharmacogenomic variation on proximal outcomes was significantly greater than that for distal outcomes. This generalises previous observations made in drug-gene guidelines about strong genetic effects on pharmacokinetics translating into weaker or unclear effects on clinical outcomes (Bousman et al., 2023b; Duarte et al., 2024). The difference in effect sizes between proximal and distal outcomes was consistent in all sensitivity analyses we made on each of the most common gene-drug subgroups in the psychiatric literature and significant in half of them, suggesting that our results are not driven by heterogeneity in the drugs or enzyme systems captured by our broad literature review. Furthermore, our analysis provides a rationale for the lack of pharmacodynamic primary evidence observed in psychiatric pharmacogenomic guidelines (Bousman et al., 2020), and supports that pharmacogenomic studies might see benefits in explicitly assessing how their phenotypes map to a “proximal-distal continuum” (Brenner et al., 1995) of genetic effects. Ideally, studies should also consider multiple outcomes relevant to clinical practice when possible, as recommended previously (Guchelaar et al., 2025).

Currently, the translation of pharmacogenomics research to clinical recommendations is primarily informed by strength of evidence ratings written into drug-gene guidelines. As an example, the five-step variant scoring system employed by PharmGKB to standardise reports includes evidence ratings by phenotype, *p*-value, cohort size, study type, and effect size (Whirl-Carrillo et al., 2021). In this calculation, the “phenotype” criteria gives a larger score to studies of drug efficacy, toxicity or dosage, while the “effect size” category increases the weight of an association if a threshold of magnitude is passed (OR >= 2, OR =< 0.5). Our results suggest that these two ratings might effectively have opposing effects, as proximal associations tend to report larger effects (pooled OR = 2.4) more commonly than distal outcomes (pooled OR = 1.6). Therefore, as the size of the pharmacogenomics literature increases and the routine access of clinicians to genomic information draws near, specific pharmacogenomic weighting rules for different phenotypes or outcome classes might need to be added to the present scheme. As there is already an explicit aim of ensuring that analyses of clinically meaningful (often distal) outcomes form the basis of drug-gene guidelines when available, such a modification would ensure a fairer evaluation of their results on the evidence assessment process.

We also investigated whether proximal-distal outcome differences in effect sizes existed across the functional spectrum of the enzymes included in our review. In these analyses we noted the distinction between estimates for proximal and distal outcomes were less consistent than in the primary analysis, with differences apparent only across the slower metabolism phenotypes (i.e., intermediate and poor). In particular, studies of poor versus normal metabolisers showed significantly larger effects than other atypical metabolism phenotypes, albeit only across proximal outcomes (**Figure 3**). These results suggest that greater care might be needed for defining metabolism phenotypes, particularly for those reflecting increased function. Furthermore, the fact that effect size estimates from ultra-rapid metabolisers versus normal metabolisers mostly overlap those of rapid and intermediate metabolisers for both proximal and distal outcomes supports that this phenotype might be particularly ill-defined in most studies. To this effect, it has already been shown that “normal” metabolisers of several enzymes have higher variability in pharmacokinetic measures when compared to atypical metabolisers identified by genetic testing, which might be attributed to the existence of functional alleles not currently assessed in pharmacogenomic studies (Lauschke et al., 2024).

Our findings have applications beyond this study. First, we provide estimates for the expected effect sizes of psychiatric pharmacogenomics analyses, based on the type of phenotype investigated. These can inform data collection efforts for future research. As an illustration, our results suggest that if per-group sample sizes are collected aiming for an appropriately-powered analysis of a proximal phenotype (e.g. drug metabolism), they are likely to be substantially underpowered for assessing a distal outcome (e.g. treatment response). Second, a retrospective analysis of our dataset using outcome-specific calculations for 80% statistical power (**Figure 4**), shows that most analyses included in our review may indeed not reach this power threshold (78.7% proximal, 84.3% distal). This suggests that barriers to participant recruitment, and potentially the availability of research funding, remain a limitation to psychiatric pharmacogenomics work (Pardiñas et al., 2021). Motivated by this finding, we conducted an estimation of the signal-to-noise ratio of past research in psychiatric pharmacogenomics using our collected studies, following the general procedure in van Zwet and Gelman (2022). The derived estimates allow for a Bayesian re-evaluation of the test statistics of comparable studies, accounting for the inflation in effect sizes that is often due to “Winner’s curse” (Zöllner and Pritchard, 2007). We provide a worked example of this methodology in the **Supplementary Material**, and implement all the relevant formulae to reproduce or replicate our calculations in a dedicated section of our Shiny app.

One of the strengths of this research is the large number of studies and effects included in the meta-analysis. This was attained through broad but well-defined inclusion criteria, namely all analyses investigating pharmacogenomic variation in cytochrome P450 (CYP) genes in participants taking psychiatric drugs. Second, based on calls to improve transparency and reproducibility in meta-analytic research (Ahern et al., 2021), we developed a companion dashboard in R Shiny that enables users to browse, filter, and download the data used in the present meta-analysis. The application also allows users to perform exploratory meta-analyses of subgroups of the data, as well as power calculations based on these results. In such a rapidly evolving field, this is not intended to substitute future efforts to compile or curate the literature but does allow users to quickly assess reported effect sizes for arbitrary combinations of drugs, enzymes, and/or outcomes that might be of interest.

A primary limitation of this research is the lack of studies identified from African and South American nations, excluding Brazil. Therefore, the results from this meta-analysis may not be generalisable to countries and participants in these locations. This needs to be taken into account as pharmacogenomic variation is known to differ across populations, in some instances quite dramatically (e.g., CYP3A4/5; Masimirembwa et al., 2014). However, consistent with a previous review (Popejoy, 2019), many studies did not report detailed ancestry information and so we were unable to statistically account for this. Another limitation derived from the assessed papers is that, while most studies used up-to-date nomenclature and metabolism phenotype definitions, several used criteria which are currently outdated. This has been corrected wherever possible, but it has not been feasible in all cases. Methodologically, visual inspection of funnel plots from the meta-analysis and results from the adapted Egger’s regression test are suggestive of funnel plot asymmetry. This has sometimes been argued as a consequence of publication bias and the inclusion of underpowered studies, but more complex factors could be at play (Afonso et al., 2024). Therefore, we made efforts to guard against potential biases in our effect size estimations by using correlated and hierarchical effects models with robust variance structures, as is best practice for datasets with potentially complex dependencies. A final limitation is that it is possible that measurement issues could partly explain our findings. Proximal outcomes such as drug pharmacokinetics and clearance can be physically measured and are thus generally easier to quantify than distal outcomes such as symptom severity. While ADRs and other distal clinical events (i.e. death) might also be easily and reliably assessed, if instruments of lower psychometric resolution are more common in distal outcomes research, this would also dilute genetic effects (Sluis et al., 2010). We are unable to account for this in our analysis as we do not have good estimates of the reliability or measurement error of all instruments involved, so our results should be considered with this caveat.

In summary, we found evidence of substantial variability in the magnitude of effect sizes reported by psychiatric pharmacogenomic studies, which we could relate to a simple phenotype classification involving either “proximal” or “distal” outcomes. We found evidence that this effect size disparity is apparent even when explicitly accounting for metabolism phenotype and is particularly pronounced for variation conferring slower enzyme activity. We have quantified these differences between proximal and distal effect sizes and provide ways in which future study design can be improved, for example via power calculations based on the results we compiled throughout our literature review. These findings may also have relevance for pharmacogenomics consortia, and future efforts focused on other drugs and disciplines of study might support new evaluations of the strength of evidence behind genotype-guided pharmacogenomic recommendations.

## Supporting information

Supplementary File (Methods and Results)

## Data Availability

All data and scripts used in the present study is available via the github repository. Data may also be accessed and downloaded via the companion shiny app.

https://locksk.shinyapps.io/pgx-effect-sizes/

https://github.com/locksk/meta-analysis-pgx

https://locksk.github.io/pgx-effects/

## Acknowledgements

We would like to thank Peter A. Holmans for critical feedback on the manuscript.

