## Supplementary File (Methods and Results) for "Near, far, wherever you are: Phenotype-related variation in pharmacogenomic effect sizes across the psychiatric drug literature"

### Supplementary Methods

#### Literature Search Terms

Literature searches were conducted across PUBMED, Cochrane, and SCOPUS on November 5^th^, 2024. Exact search varied by database; however, the following terms and Boolean operators were used to search across titles:

Pharmacogenomics keyword AND

("pharmacogen*" OR "CYP" OR "CYP2C19" OR "CYP2D6" OR "CYP1A2" OR "CYP3A4" OR "CYP3A5" OR "CYP2C9" OR "cytochrome P450" OR "genotype" OR "allele" OR "star allele" OR "metabol*" OR "metabol* status" OR "metabol* phenotype" OR "poor metabol*" OR "intermediate metabol*" OR "extensive metabol*" OR "normal metabol*" OR "rapid metabol*" OR "ultrarapid metabol*" OR "ultra-rapid metabol*" OR "activity score" OR "activity" OR "increased function" OR "decreased function")

Medication keyword AND

("psychiatric" OR "drugs" OR "medication" OR "clozapine" OR "risperidone" OR "quetiapine" OR "olanzapine" OR "aripiprazole" OR "haloperidol" OR "antipsychotic" OR "atypical antipsychotic" OR "typical antipsychotic" OR "neuroleptic" OR "sertraline" OR "escitalopram" OR "amitriptyline" OR "nortriptyline" OR "paroxetine" OR "venlafaxine" OR "fluvoxamine" OR "fluoxetine" OR "antidepressants" OR "SSRIs")

Outcome keyword AND

("metabolic ratio" OR "concentration" OR "levels" OR "serum" OR "plasma" OR "moiety" OR "metabolite" OR "clearance" OR "AUC" OR "half life" OR "C/D" OR "dose" OR "dose adjustment" OR "sympt*" OR "improve*" OR "increas*" OR "decreas*" OR "reduc*" OR "sever*" OR "tolerability" OR "response" OR "remission" OR "discontinuation" OR "therapeutic outcome" OR "treatment outcome" OR "therapeutic response" OR "failure" OR "symptom" OR "efficacy" OR "compliance" OR "adherence" OR "extra-pyramidal" OR "extra pyramidal" OR "side effects" OR "adverse effects" OR "adverse drug reaction" OR "ADR" OR "adverse event" OR "QTc" OR "prolactin" OR "side effect" OR "tardive" OR "weight gain" OR "switch" OR "sedation" OR "drop out" OR "score" OR "suicid*" OR "resistance" OR "refractory" OR "responsive" OR "hospital*")

AND NOT Entry type keyword

("Guidelines" OR "Guideline" OR "Review" OR "guidelines" OR "guideline" OR "review" OR "case study" OR "case series" OR "case report")

In the PUBMED search only, we filtered to studies conducted on humans. Any studies conducted *in vitro* or on non-human animals that were identified from other databases were excluded during the later screening stages.

#### Effect size conversions

Across the included studies, results were reported using a range of statistics such as measures of central tendency, observations, and those from inferential tests. These were used to calculate the standardised mean difference of the pharmacogenomic effect as outlined in Supplementary Table 1. When SMD was calculated based on descriptive statistics or observations, we used a minimum per-group sample size of N = 5 (Curtis et al. 2015), and per-analysis sample size of N = 25 for regression models (Jenkins and Quintana-Ascencio 2020).

| **Reported Effect Size** | **Reported Variance** | **Conversions** | **Required Effect Size (etc)** | **package::function()** |
| --- | --- | --- | --- | --- |
| Mean | Standard deviation | *NA* | Mean and Standard deviation | *esc::esc_mean_sd()* |
|  | Confidence intervals | Confidence Intervals to Standard Error to Standard Deviation.^1^ |  |  |
|  | Standard Errors | Confidence Intervals to Standard Error.^2^ |  |  |
| Median | Interquartile range, Min, Max | Median and IQR and Range to Mean^3^ and Standard Deviation^4^ |  |  |
|  | Range | Median and Range to Mean^5^ and Standard Deviation^6^ |  |  |
|  | Interquartile range | Median and IQR to Mean^7^ and Standard Deviation^8^ |  |  |
| χ^2^ | *NA* | *NA* | χ^2^/*p N* | *esc::esc_2x2()* |
| OR | Confidence intervals | Confidence Intervals to Standard Error^9^ | OR and Standard error | *esc::convert_or2d()* |
| β | SE | Beta and SE to *t*^10^ | *t, p,* total N | *esc::esc_t()* |
| *t* | *NA* | *NA* | *t, p,* total N |  |

Supplementary Table 1. Overview of equations used to calculate effect sizes from reported data in the present study.

##### Measures of Central Tendency

Means reported alongside standard deviations (SD), standard errors (SE), or confidence intervals (CIs) can be converted to standardised mean difference with the *esc::esc_mean_sd()* function. The mean and standard deviation do not require further conversion. Standard errors and confidence intervals need to be transformed to a standard deviation prior to running the function.

95% confidence intervals for a mean can be converted to standard errors as shown below. The denominator is taken from the standard normal distribution, with a value matching the confidence interval being transformed, e.g. 3.92 is used for 95% confidence intervals.

1. $SE= \frac{CI \left( Upper \right)-CI (Lower)}{3.92}$

Where groups have small sample sizes (N), the confidence intervals should have been sampled from the *t* distribution, and not the normal distribution. At low values (N< 30), the *t* distribution and the normal distribution differ, and so when the group N is lower than 30 the denominator is instead replaced with the inverse of the *t* distribution for the given sample size (Higgins et al. 2024).

Standard error is converted to standard deviation as follows.

2. $SD= SE \times\surd N$

Medians were also reported alongside interquartile ranges and/or ranges. We estimated means and standard deviations using the following equations (Wan et al. 2014), as these allow for better estimates at low sample sizes than traditional methods (Hozo et al. 2005; Bland 2014).

When median, IQR, Min, Max are available, mean and standard deviation are calculated as so:

3. $Mean=\frac{Min+\left( 2\times Q1 \right)+\left( 2\times Median \right)+\left( 2\times Q3 \right)+Max}{8}$
4. $SD=\frac{Max-Min}{4\times qnorm\left( \left( \frac{N-0.375}{N+0.25} \right),0,1 \right)}+\frac{Q3-Q1}{4\times qnorm\left( \left( \frac{0.75\times N-0.125}{N+0.25} \right),0,1 \right)}$

When only the median and range are available, the formulae become:

5. $Mean=\frac{Min+\left( 2\times Median \right)+Max}{4}$
6. $SD=\frac{Max-Min}{2\times qnorm\left( \left( \frac{N-0.375}{N+0.25} \right),0,1 \right)}$

When only the median and IQR are available, the formulae become:

7. $Mean= \frac{Q1+Median+Q3}{3}$
8. $SD=\frac{Q3-Q1}{2\times qnorm\left( \left( \frac{0.75\times N-0.125}{N+0.25} \right),0,1 \right)}$

Where intervention studies report statistics as a change between baseline and a given time point, we calculated the SMD at each time point separately. We did not combine SMDs calculated at a single time point with those representing a change over time, following the recommendations in the Cochrane handbook (Higgins et al. 2024). When single time point measures were not available to calculate SMDs, the reported change-from-baseline statistics were excluded from the analysis.

##### Observations

Where observational data were reported, the SMD was calculated using *esc::esc_2x2()*. In instances where a cell count in the 2x2 table was 0, we performed a Haldane-Anscombe correction by adding 0.5 to all counts in that table to allow the SMD to be calculated (Weber et al. 2020).

##### Measures of Effect (Ratio Measures)

Odds ratios (OR) are often reported with confidence intervals. The latter must be converted into standard errors prior to estimating the SMD. The equation is the same as above, however the natural log of both the upper and lower confidence intervals is used instead. There is no need to sample from the *t-*distribution at low sample sizes. Where variables were log-transformed prior to use in the model, the OR was back-transformed.

9. $SE= \frac{log(CI \left( Upper) \right)-log(CI \left( Lower \right))}{3.92}$

##### Measures of Effect (Absolute Measures)

Many linear regression analyses report beta coefficients, often alongside standard errors. These cannot be directly converted to SMD. However, they can be converted to a *t* statistic which can then be converted to SMD. Where variables were log-transformed prior to use in the model, the β was back-transformed.

10. $t= \beta\times SE$

The *t* statistic can be directly converted to an SMD estimate by using the *t* statistic, its *p* value and the total number of participants analysed using *esc:: esc_t()*.

##### Metabolic Ratios

Metabolic ratios in the pharmacokinetic literature are reported in one of two ways:

i) Metabolic Ratio = drug / metabolite
ii) Metabolic Ratio = metabolite / drug

The interpretation of these is different, with higher values reflecting slower metabolism in the first case, and faster metabolism in the second case. Although the interpretation is slightly less intuitive, most metabolic ratios are reported with the parent drug divided by the metabolite. Therefore, all other metabolic ratios have been converted so that they are equivalent to this standard. This method varies slightly, dependent on whether results are given as mean, $\beta$, or ORs (see equations 11 – 13). The same method was applied to their standard deviations, standard errors, and confidence intervals, respectively.

11. $Mean\frac{D}{M}=exp(\log\left( Mean\frac{M}{D} \right)\times-1)$
12. $\beta\frac{D}{M}=\beta\frac{M}{D}\times-1$
13. $OR\frac{D}{M}=exp(log(OR\frac{M}{D})\times-1))$

##### Joining Groups

There are also instances where we wanted to calculate metabolism phenotypes based on per-genotype means/SDs or combine metabolism phenotypes if they have been inappropriately inferred. To do this we have followed Cochrane guidance (Higgins et al. 2024) using equations 14 and 15.

14. $Mean= \frac{N_{1}M_{1}+ N_{2}M_{2}}{N_{1}+N_{2}}$
15. $SD=\sqrt{\frac{{(N}_{1}-1){SD}_{1}^{2}+{(N}_{2}-1){SD}_{2}^{2}+\frac{N_{1}N_{2}}{N_{1}+ N_{2}}(M_{1}^{2}+M_{2}^{2}- 2M_{1}M_{2})}{N_{1}+ N_{2}-1}}$

#### Pharmacogenomic Alleles and Metabolism Phenotypes

We used information from PharmGKB Gene-Specific Information tables to define allele functions and metabolism phenotypes. Where genetic information was reported as SNPs, we used information from PyPGx to identify the genetic variants that characterise PharmGKB alleles. Analyses were excluded where CYP metabolism phenotypes were inferred using non-genetic approaches, or where SNPs were not aligned with pharmacogenomic star allele definitions. We were unable to resolve metabolism phenotypes for 420 analyses.

When reviewing the SNP-to-allele table for CYP2D6, we noted many studies in our meta-analysis called CYP2D6*10 based on a single SNP (rs1065852), despite that SNP being found in multiple CYP2D6 star alleles. This definition is a simplification of the allelic landscape of CYP2D6 but has been noted before as common throughout the pharmacogenomics literature (Nofziger et al. 2020). In an effort to best standardise pharmacogenomic allele calling across our meta-analysis, we have chosen to assign CYP2D6*10 alleles, and where possible CYP2D6 metabolism phenotypes, using solely rs1065852. We acknowledge this is a potential limitation of our study, caused by a common practice that should be addressed by future studies. The retrieval date of the PharmGKB/PyPGx tables was 17/07/2025.

##### CYP1A2

Gene-specific information tables are not available for this gene; therefore, allele definitions and functions were taken from the PyPGx information tables. There is no current consensus regarding the mapping of star alleles to metabolism phenotypes for CYP1A2 (Fekete et al. 2022; Kuhn et al. 2025), therefore pharmacogenomic information has been in the format of star allele diplotypes.

| **SNP** | **Allele** | **Function** | **Source** |
| --- | --- | --- | --- |
| - | *1A | Normal Function | PyPGx |
| rs2069514 (-2964G>A; -3860G>A) | *1C | Decreased Function | PyPGx |
| rs762551 (-173C>A; 163C>A) | *1F | Increased function | PyPGx |

Supplementary Table 2. Functions of CYP1A2 SNPs and pharmacogenomic alleles referenced in the studies curated for this meta-analysis.

##### CYP2B6

PharmGKB gene-specific information tables are available and have been used in the present study. Metabolism phenotypes were determined based on the functionality of alleles in the diplotype.

| **SNPs** | **Allele** | **Function** | **Source** |
| --- | --- | --- | --- |
| - | *1 | Normal function | PyPGx, PharmGKB |
| rs3745274, rs2279343 | *6 | Decreased function | PyPGx, PharmGKB |
| rs3745274 | *9 | Decreased function | PyPGx, PharmGKB |

Supplementary Table 3. Functions of CYP2B6 SNPs and pharmacogenomic alleles referenced in the studies curated for this meta-analysis.

| **Diplotype** | **Metabolism Phenotype** | **Source** |
| --- | --- | --- |
| *1/*1 | Normal Metabolizer | PyPGx, PharmGKB |
| *1/*6 | Intermediate Metabolizer | PyPGx, PharmGKB |
| *6/*6 | Poor Metabolizer | PyPGx, PharmGKB |
| *6/*9 | Poor Metabolizer | PyPGx, PharmGKB |

Supplementary Table 4. Metabolism phenotypes for CYP2B6 diplotypes referenced in the studies curated for this meta-analysis.

##### CYP2C9

PharmGKB gene-specific information tables were available and have been used in the present study. Metabolism phenotypes were determined based on the functionality of alleles in the diplotype, and by extension the total activity score.

- Poor Metaboliser is assigned to those with activity scores 0 <= score < 1.
- Intermediate Metaboliser is assigned to those with activity scores 1 <= score < 2.
- Normal Metaboliser is assigned to those with activity scores 2 == score.

| **Allele** | **Function** | **Activity Score** | **Source** |
| --- | --- | --- | --- |
| *1 | Normal function | 1 | PyPGx, PharmGKB |
| *2 | Decreased function | 0.5 | PyPGx, PharmGKB |
| *3 | Null function | 0 | PyPGx, PharmGKB |

Supplementary Table 5. Functions of CYP2C9 pharmacogenomic alleles referenced in the studies curated for this meta-analysis.

| **Diplotype** | **Metabolism Phenotype** | **Total Activity Score** | **Source** |
| --- | --- | --- | --- |
| *1/*1 | Normal metaboliser | 2 | PyPGx, PharmGKB |
| *1/*2 | Intermediate metaboliser | 1.5 | PyPGx, PharmGKB |
| *1/*3 | Intermediate metaboliser | 1 | PyPGx, PharmGKB |
| *2/*2 | Intermediate metaboliser | 1 | PyPGx, PharmGKB |
| *2/*3 | Poor metaboliser | 0.5 | PyPGx, PharmGKB |
| *3/*3 | Poor metaboliser | 0 | PyPGx, PharmGKB |

Supplementary Table 6. Metabolism phenotypes for CYP2C9 diplotypes referenced in the studies curated for this meta-analysis.

##### CYP2C19

PharmGKB gene-specific information tables were available and have been used in the present study. Metabolism phenotypes were determined based on the functionality of alleles in the diplotype.

| **SNPs** | **Allele** | **Function** | **Source** |
| --- | --- | --- | --- |
| **-** | *1 | Normal function | PyPGx, PharmGKB |
| rs4244285, rs12769205 | *2 | Null function | PyPGx, PharmGKB |
| rs4986893 | *3 | Null function | PyPGx, PharmGKB |
|  | *4 | Null function | PyPGx, PharmGKB |
|  | *5 | Null function | PyPGx, PharmGKB |
|  | *6 | Null function | PyPGx, PharmGKB |
| rs12248560 | *17 | Increased function | PyPGx, PharmGKB |

Supplementary Table 7. Functions of CYP2C19 SNPs and pharmacogenomic alleles referenced in the studies curated for this meta-analysis.

| **Diplotype** | **Metabolism Phenotype** | **Source** |
| --- | --- | --- |
| *1/*1 | Normal Metabolizer | PyPGx, PharmGKB |
| *1/*2 | Intermediate Metabolizer | PyPGx, PharmGKB |
| *1/*3 | Intermediate Metabolizer | PyPGx, PharmGKB |
| *1/*4 | Intermediate Metabolizer | PyPGx, PharmGKB |
| *1/*5 | Intermediate Metabolizer | PyPGx, PharmGKB |
| *1/*6 | Intermediate Metabolizer | PyPGx, PharmGKB |
| *1/*17 | Rapid Metaboliser | PyPGx, PharmGKB |
| *2/*2 | Poor Metabolizer | PyPGx, PharmGKB |
| *2/*3 | Poor Metabolizer | PyPGx, PharmGKB |
| *2/*4 | Poor Metabolizer | PyPGx, PharmGKB |
| *2/*17 | Intermediate Metabolizer | PyPGx, PharmGKB |
| *3/*3 | Poor Metabolizer | PyPGx, PharmGKB |
| *17/*17 | Ultrarapid Metaboliser | PyPGx, PharmGKB |

Supplementary Table 8. Metabolism phenotypes for CYP2C19 diplotypes referenced in the studies curated for this meta-analysis.

##### CYP2D6

PharmGKB gene-specific information tables were available and have been used in the present study Metabolism phenotypes are determined based on the functionality of alleles, via their activity scores, in the diplotype as follows:

- Poor Metaboliser is assigned to those with activity scores 0 <= score < 0.25.
- Intermediate Metaboliser is assigned to those with activity scores 0.25 <= score < 1.25.
- Normal Metaboliser is assigned to those with activity scores 1.25 <= score < 2.5.
- Ultrarapid Metaboliser is assigned to those with activity scores 2.5 <= score.

| **SNP** | **Allele** | **Activity Score** | **Function** | **Source** |
| --- | --- | --- | --- | --- |
| - | *1 | 1 | Normal Function | PyPGx, PharmGKB |
| - | *1x2 | 2.0 | Increased function | PharmGKB |
|  | *2 | 1.0 | Normal function | PyPGx, PharmGKB |
| rs35742686 | *3 | 0 | Null Function | PyPGx, PharmGKB |
| rs3892097 (1846G>A) | *4 | 0 | Null Function | PyPGx, PharmGKB |
|  | *5 | 0 | Null Function | PyPGx, PharmGKB |
|  | *6 | 0 | Null Function | PyPGx, PharmGKB |
|  | *7 | 0 | Null Function | PyPGx, PharmGKB |
|  | *8 | 0 | Null Function | PyPGx, PharmGKB |
| rs1065852 | *10 | 0.25 | Decreased | PyPGx, PharmGKB |
| rs5030865^†^ | *14 | 0.5 | Decreased | PyPGx, PharmGKB |
| ^††^ | *41 | 0.25 | Decreased | PharmGKB |

Supplementary Table 9. Functions of CYP2D6 SNPs and pharmacogenomic alleles referenced in the studies curated for this meta-analysis. ^†^T allele reflects CYP2D6*14, whereas A allele at this location reflects CYP2D6*8. ^††^Activity scores for *41 conflict between PyPGx (0.5) and PharmGKB (0.25). PharmGKB is the newest so was used for the present study.

| **Diplotype** | **Total Activity Score** | **Metabolism Phenotype** | **Source** |
| --- | --- | --- | --- |
| *1/*1 | 2 | Normal Metaboliser | PharmGKB |
| *1/*2 | 2 | Normal Metaboliser | PharmGKB |
| *1/*3 | 1 | Intermediate Metaboliser | PharmGKB |
| *1/*4 | 1 | Intermediate Metaboliser | PharmGKB |
| *1/*5 | 1 | Intermediate Metaboliser | PharmGKB |
| *1/*6 | 1 | Intermediate Metaboliser | PharmGKB |
| *1/*7 | 1 | Intermediate Metaboliser | PharmGKB |
| *1/*8 | 1 | Intermediate Metaboliser | PharmGKB |
| *1/*10 | 1.25 | Normal Metaboliser | PharmGKB |
| *1/*41 | 1.25 | Normal Metaboliser | PharmGKB |
| *1/(*1x2) | 3 | Ultrarapid metaboliser | PharmGKB |
| *1/(*1xN) | > 2 | Ultrarapid metaboliser | PharmGKB |
| *1/(*2xN) | > 2 | Ultrarapid metaboliser | PharmGKB |
| *2/*2 | 2 | Normal Metaboliser | PharmGKB |
| *2/*3 | 1 | Intermediate Metaboliser | PharmGKB |
| *2/*5 | 1 | Intermediate Metaboliser | PharmGKB |
| *2/*10 | 1.25 | Normal Metaboliser | PharmGKB |
| *2/*41 | 1.25 | Normal Metaboliser | PharmGKB |
| *3/*4 | 0 | Poor Metaboliser | PharmGKB |
| *3/*5 | 0 | Poor Metaboliser | PharmGKB |
| *4/*4 | 0 | Poor Metaboliser | PharmGKB |
| *4/*5 | 0 | Poor Metaboliser | PharmGKB |
| *4/*6 | 0 | Poor Metaboliser | PharmGKB |
| *4/*10 | 0.25 | Intermediate Metaboliser | PharmGKB |
| *4/*14 | 0.5 | Intermediate Metaboliser | PharmGKB |
| (2x*1)/*4 | 2 | Normal Metaboliser | PharmGKB |
| *5/*6 | 0 | Poor Metaboliser | PharmGKB |
| *5/*10 | 0.25 | Intermediate Metaboliser | PharmGKB |
| *6/*6 | 0 | Poor Metaboliser | PharmGKB |
| *10/*10 | 0.5 | Intermediate Metaboliser | PharmGKB |
| *10/*41 | 0.5 | Intermediate Metaboliser | PharmGKB |

Supplementary Table 10. Metabolism phenotypes for CYP2D6 diplotypes referenced in the studies curated for this meta-analysis.

##### CYP3A4

PharmGKB gene-specific information tables were available and have been used in the present study. There is no information regarding genotype to metabolism phenotype conversion and so genetic information was retained in its original format. are determined based on the functionality of alleles in the diplotype.

|  | **Allele** | **Function** | **Source** |
| --- | --- | --- | --- |
|  | *1 | Normal Function | PyPGx |
| rs2740574 | *1B | Unknown Function | PharmGKB |
|  | *22 | Unknown Function | PyPGx |

Supplementary Table 11. Functions of CYP3A4 SNPs and pharmacogenomic alleles referenced in the studies curated for this meta-analysis.

##### CYP3A5

PharmGKB gene-specific information tables were available and have been used in the present study. Metabolism phenotypes are determined based on the functionality of alleles in the diplotype.

| **Allele** | **Function** | **Source** |
| --- | --- | --- |
| *1 | Normal function | PyPGx |
| *3 | Null function | PyPGx |
| *6 | Null function | PyPGx |

Supplementary Table 12. Functions of CYP3A5 pharmacogenomic alleles referenced in the studies curated for this meta-analysis.

| **Diplotype** | **Metabolism Phenotype** | **Source** |
| --- | --- | --- |
| *1/*1 | Normal Metaboliser | PyPGx |
| *1/*3 | Intermediate Metaboliser | PyPGx |
| *1/*6 | Intermediate Metaboliser | PyPGx |
| *3/*3 | Poor Metaboliser | PyPGx |
| *3/*6 | Poor Metaboliser | PyPGx |
| *6/*6 | Poor Metaboliser | PyPGx |

Supplementary Table 13. Metabolism phenotypes for CYP3A5 diplotypes referenced in the studies curated for this meta-analysis.

### Supplementary Results

#### Addressing “winner’s curse” in psychiatric pharmacogenomic studies.

The dataset compiled for this work can also be used to evaluate the results of psychiatric pharmacogenomic studies through a Bayesian perspective. Following a general procedure described by van Zwet and Gelman (2022), information from a corpus of existing studies can be used to estimate the degree of potential inflation in a novel result, a phenomenon that often leads to poor replicability known as “winner’s curse” (Zöllner and Pritchard 2007; Ioannidis 2008; van Zwet and Cator 2021). Briefly, this requires modelling a prior distribution of signal-to-noise ratios using reported z-scores. Then, for any new result in the form of regression coefficients and variances, this prior can be used to derive a posterior expectation of unbiased effects (van Zwet and Gelman 2022), assuming the new study is exchangeable with other studies in the corpus (i.e. fits a set of inclusion criteria, in our case focusing on psychiatric medications, CYP metaboliser phenotypes, and an outcome that can be categorised as either “proximal” or “distal”). Such a Bayesian reanalysis of frequentist test results has similarities with regression regularisation procedures (Bickel et al. 2006), but without the need for raw data, making it particularly useful to critically assess published evidence when undertaking translational research (Kraft 2008; van Zwet et al. 2021).

We exemplify this procedure by re-analysing a result from a large-scale pharmacogenomic study of 9,563 antidepressant users (escitalopram, citalopram, sertraline or amitriptyline) published after the end date of our literature search (Kariis et al. 2025). The analysis of this cohort showed CYP2C19 poor metabolisers to have an increased risk of reporting adverse effects, a result consistent with existing pharmacogenomic drug-gene guidelines (Brouwer et al. 2022; Bousman et al. 2023). The reported effect for this association is OR=1.485 (95%CI=1.087–2.038). From our corpus of results of distal outcomes without filtering by drug or gene (107 studies, 930 effect sizes), we modelled a two-component mixture of zero-mean normal distributions using the *flexmix* R package (Leisch 2004), using the study ID as a grouping factor. This distribution is a prior for the signal-to-noise ratio of distal outcome studies, and an equivalent prior for reports of proximal outcomes (124 studies, 1162 effect sizes) is illustrated for comparison (Supplementary Figure 1). The mixture distribution for distal outcomes had SD1=0.661 and SD2=4.015, with mixture weights W1=0.818 and W2=0.182. Using code from van Zwet and Gelman (2022), we use these parameters and the original regression betas and standard errors to estimate a posterior mean of OR=1.22 (95%CI=0.96–1.805) for the pharmacogenomic result above. This implies a reduction of the magnitude of the original estimate by a factor of 1.992 after accounting for winner’s curse. Nevertheless, the re-analysis still supports, with 93.76% probability, that CYP2C19 poor metabolism indeed increases the likelihood of reporting adverse effects in individuals prescribed certain antidepressants.

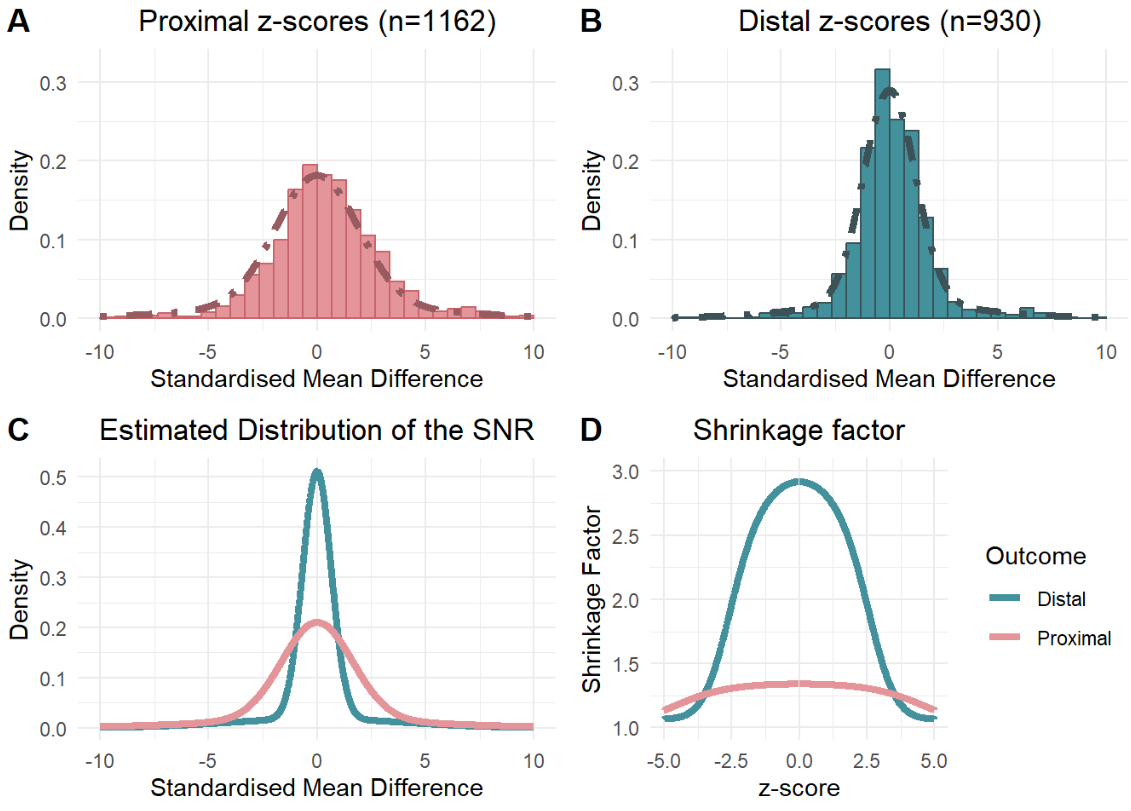

Supplementary Figure 1. Panel A and B show the distribution of Z-scores observed for proximal and distal outcomes, respectively. The line represents the fitted mixtures of two zero-mean normals for both outcomes. Panel C shows the estimated distribution of the signal-to-noise ratio (SNR) for both proximal and distal outcomes. Panel D shows variation in the shrinkage factor (the ratio of estimated to original effect sizes) across z-values for proximal and distal outcomes. For a new effect size, posterior mean estimates can be calculated by dividing it by the shrinkage factor; thus, the amount of shrinkage decreases as the Shrinkage Factor approaches 1.

#### Assessing random-effects structure.

Model structure was guided by knowledge of dependencies within the dataset, and goodness-of-fit statistics. We fit several different random effects structures allowing random intercepts for (i) Study, (ii) Study / Analysis, and (iii) Cohort / Study / Analysis. These tests were based on simplified versions of the primary model in which no variance-covariance matrix was used in place of effect size variances. Analysis of Variance suggested that model ii demonstrated better fit than model i (LRT = 6610, *p* < 0.0001). There was no significant difference between model ii, and model iii which also includes a random effect for cohort (LRT = 0, *p* = 1).

Finally, we tested a more complex random effect structure to allow the variance components at the Study level to differ by the moderator, “rating”. Overall, this was the best fit model and so was used in the primary analysis (LRT = 14.426, *p* = 0.001). However, this structure requires substantial data for its components to be properly estimated and so is not feasible for the sensitivity analyses on the gene-drug subgroups; therefore, we used a simplified random-effects structure for these analyses (i.e., ~1 | Study ID / Effect Size ID). For the secondary analysis which contained an interaction term between rating and metabolism phenotype, we followed the primary analysis, including a “rating| Study” term. This demonstrated better model fit than the simple nested structure (LRT = 8.58, *p* = 0.014), while balancing the risk for over-parametrisation that would arise with including a term for each moderator.

Supplementary Figures 2 – 7 show profile-Likelihood plots for the variance components of the primary meta-analysis, the secondary meta-analysis, and the four sensitivity analyses. The plots do not indicate that the models are over-parametrised; the restricted log-likelihood peaks at the parameter estimates and gradually decreases on either side which suggests confidence in these estimates (as opposed to flat or bimodal profiles, which could indicate lack of parameter identifiability).

Final model structures for the various analyses are described below:

i) For the primary analysis, the absolute SMD was moderated by rating (i.e., whether the outcome was proximal or distal) and included random effect terms for Study ID (allowed to vary by rating) and Effect Size ID.

rma.mv(absmd ~ 0 + rating, V = V, random = list(~ rating | Study, ~ 1 | es_id), struct = "UN", method = "REML", test = "t", dfs = 'contain’, data = all_smd, sparse = TRUE)

ii) For the sensitivity analyses, the absolute SMD was moderated by rating and included a simplified random effect term for Effect Size ID nested within Study ID (example given for Risperidone-CYP2D6 analysis).

rma.mv(absmd ~ 0 + rating, V = Vris, random = list(~ 1 | Study / es_id), method = "REML", test = "t", dfs = 'contain', data = ris_smd, sparse = TRUE)

iii) For the secondary analysis, the absolute SMD was moderated by a rating-metabolism phenotype interaction and included random effect terms for Study ID (allowed to vary by rating) and Effect Size ID.

meta.int <- rma.mv(absmd ~ 0 + rating:mp, V = V, random = list(~ rating | Study, ~ 1 | es_id), struct = c("UN"), method = "REML", test = "t", dfs = 'contain', data = meta.dat.all, sparse = TRUE)

#### Profile Likelihood plots for Assessing Model Over-parameterisation.

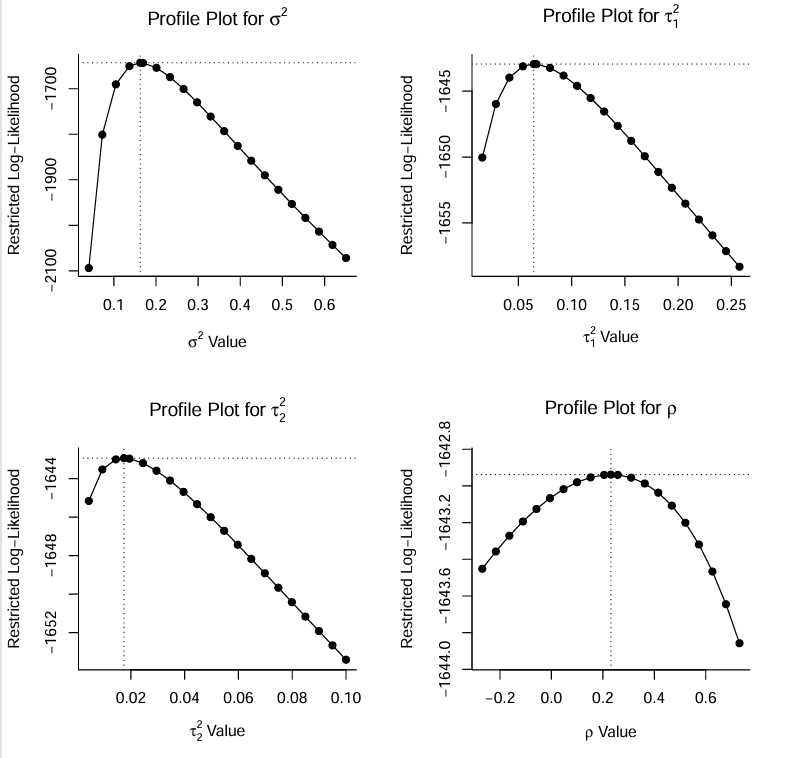

Supplementary Figure 2. Profile likelihood plots for the variance components for the primary analysis. The X axis represents variance component estimates, with the dotted line intersecting with the X axis represents the value of the variance component as calculated by the primary model. The Y axis represents the restricted log-Likelihood at each variance component estimate.

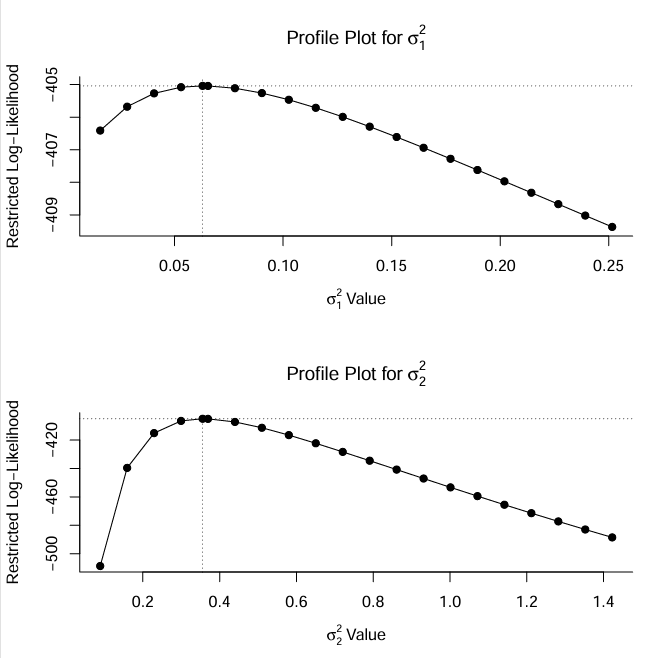

Supplementary Figure 3. Profile likelihood plots for the variance components of the sensitivity analysis (Risperidone and CYP2D6). The X axis represents variance component estimates, with the dotted line intersecting with the X axis represents the value of the variance component as calculated by the primary model. The Y axis represents the restricted log-Likelihood at each variance component estimate.

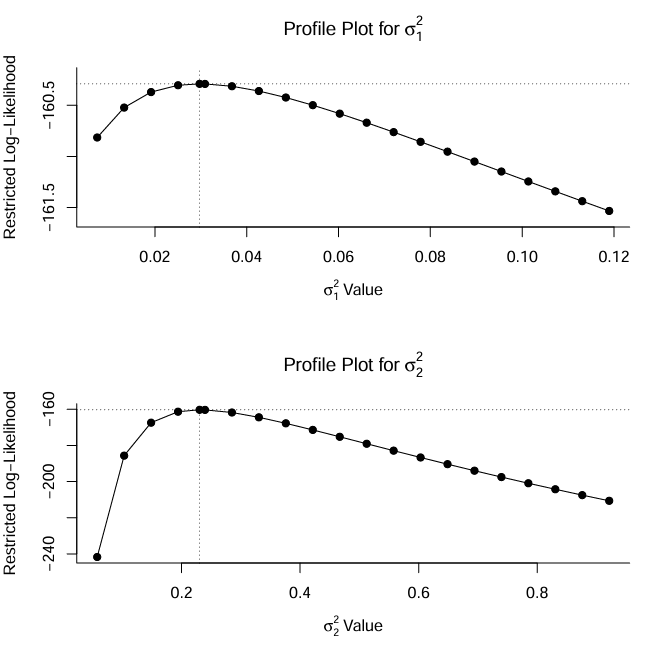

Supplementary Figure 4. Profile likelihood plots for the variance components of the sensitivity analysis (Escitalopram and CYP2C19). The X axis represents variance component estimates, with the dotted line intersecting with the X axis represents the value of the variance component as calculated by the primary model. The Y axis represents the restricted log-Likelihood at each variance component estimate.

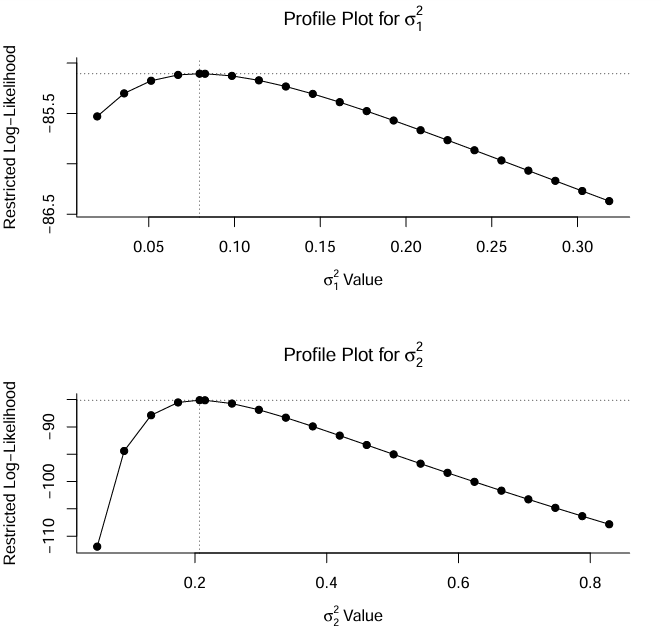

Supplementary Figure 5. Profile likelihood plots for the variance components of the sensitivity analysis (Aripiprazole and CYP2D6). The X axis represents variance component estimates, with the dotted line intersecting with the X axis represents the value of the variance component as calculated by the primary model. The Y axis represents the restricted log-Likelihood at each variance component estimate.

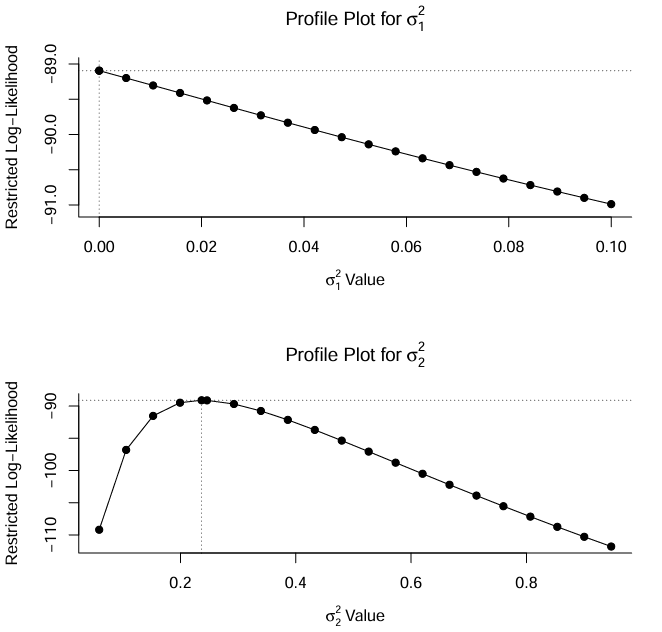

Supplementary Figure 6. Profile likelihood plots for the variance components of the sensitivity analysis (Haloperidol and CYP2D6). The X axis represents variance component estimates, with the dotted line intersecting with the X axis represents the value of the variance component as calculated by the primary model. The Y axis represents the restricted log-Likelihood at each variance component estimate.

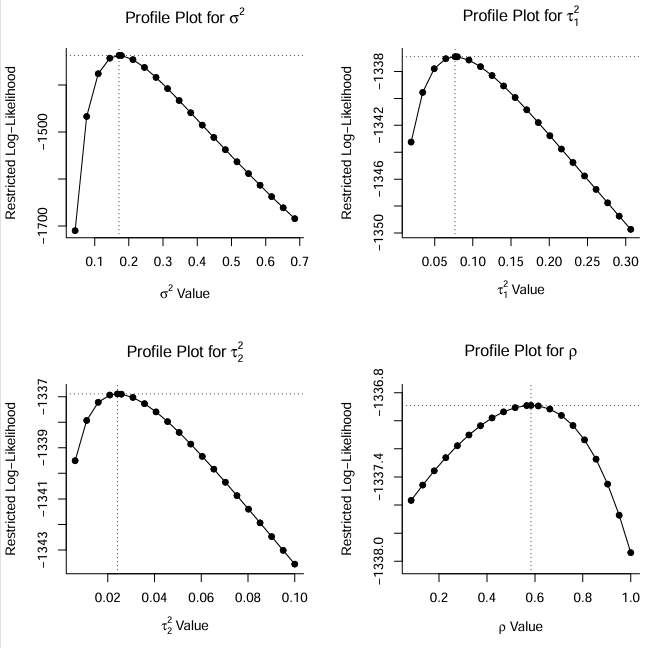

Supplementary Figure 7. Profile likelihood plots for the variance components of the secondary analysis. The X axis represents variance component estimates, with the dotted line intersecting with the X axis represents the value of the variance component as calculated by the secondary model. The Y axis represents the restricted log-Likelihood at each variance component estimate.

#### Approximating the Variance-Covariance matrix using different values of ρ.

To account for dependencies in our data, we imputed a variance-covariance matrix and use this in place of effect size variances or standard errors as the variance argument in our meta-analysis. In the absence of information regarding correlations in raw data, we used ρ = 0.6 (i.e. assuming sampling correlation of 0.6 between effects of the same study) to impute the matrix used in our primary analysis as suggested in previous work (Pustejovsky and Tipton 2022).

As a sensitivity analysis, we tested how our effect estimates were affected by assuming different sampling correlations (i.e., changing the value of ρ, from 0 to 0.95 at intervals of 0.05). We plot the estimate and confidence intervals for the difference between proximal and distal outcomes (Supplementary Figure 8), as well as examining the variance components (i.e., σ^2^, τ^2.1^, τ^2.2^ as in Supplementary Figure 9). Overall, there is little effect of changing the assumed sampling correlation, ρ, on key model coefficients or variance components.

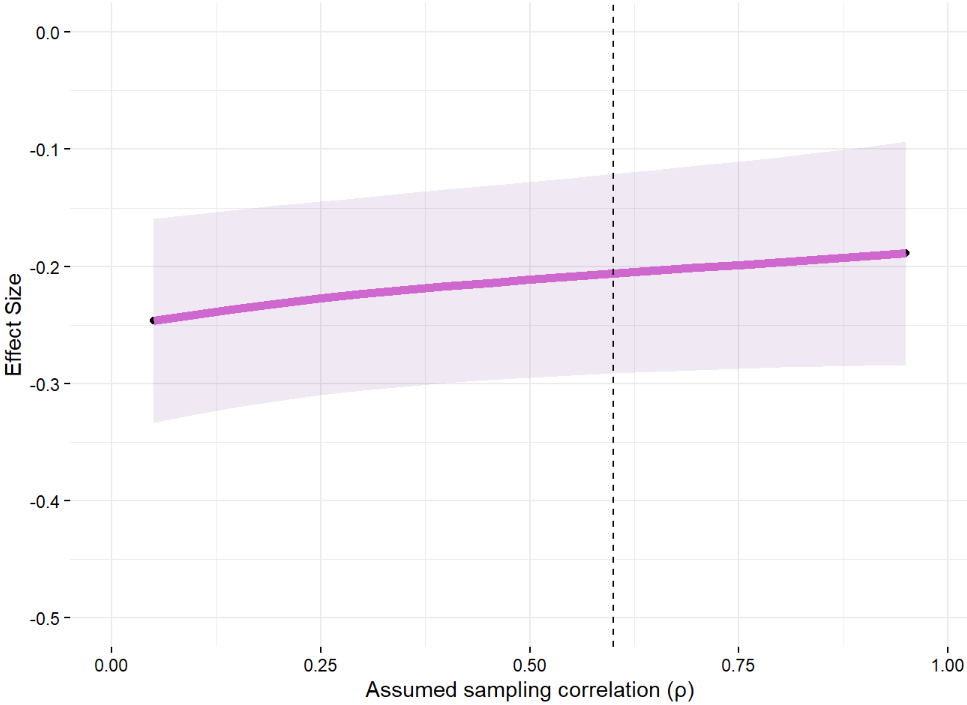

Supplementary Figure 8. Estimates for the difference between proximal and distal effect sizes are plotted at each value of the assumed constant sampling correlation (ρ) in the imputation of the variance-covariance matrix for the primary meta-analysis. The vertical dashed line represents ρ = 0.6, the default value recommended in Pustejovsky and Tipton (2022). Estimates are from an intercept model. Shaded area represents robust 95% confidence intervals around the estimates.

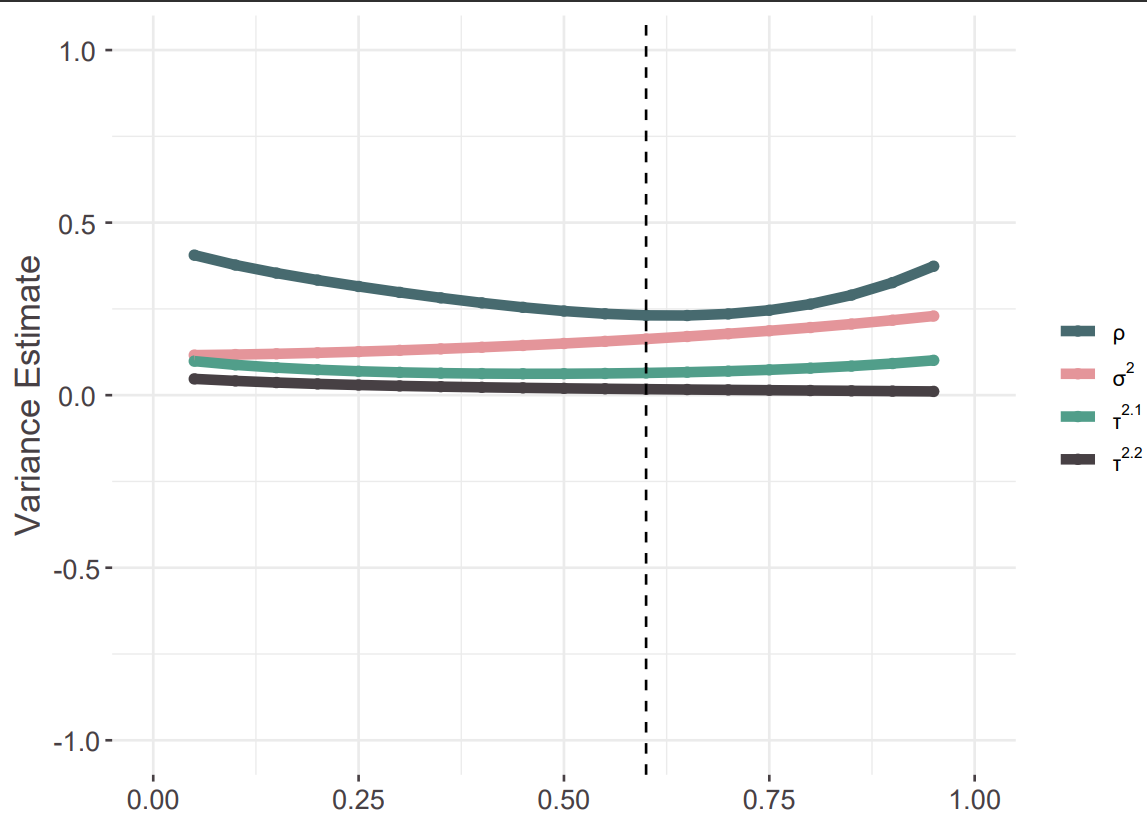

Supplementary Figure 9. Variance component estimates for within-study (σ^2^), between-study (proximal; τ^2.1^) and between-study (distal; τ^2.2^) heterogeneity from across different values of the assumed sampling correlation coefficient (ρ) in the imputation of the variance-covariance matrix for the primary meta-analysis. The vertical dashed line represents ρ = 0.6, the value recommended in Pustejovsky and Tipton (2022).

#### Assessing Risk of Bias

Funnel plots suggest a risk of small-study effects among the studies included in the primary meta-analysis (Supplementary Figure 10). This is supported by our results from an adapted version of Egger’s test (estimate = 2.32, *p* = 8x10^-8^). Our methodology fits with current best-practice for dealing with biases, including publication bias, in similar meta-analyses (Yang et al. 2024).

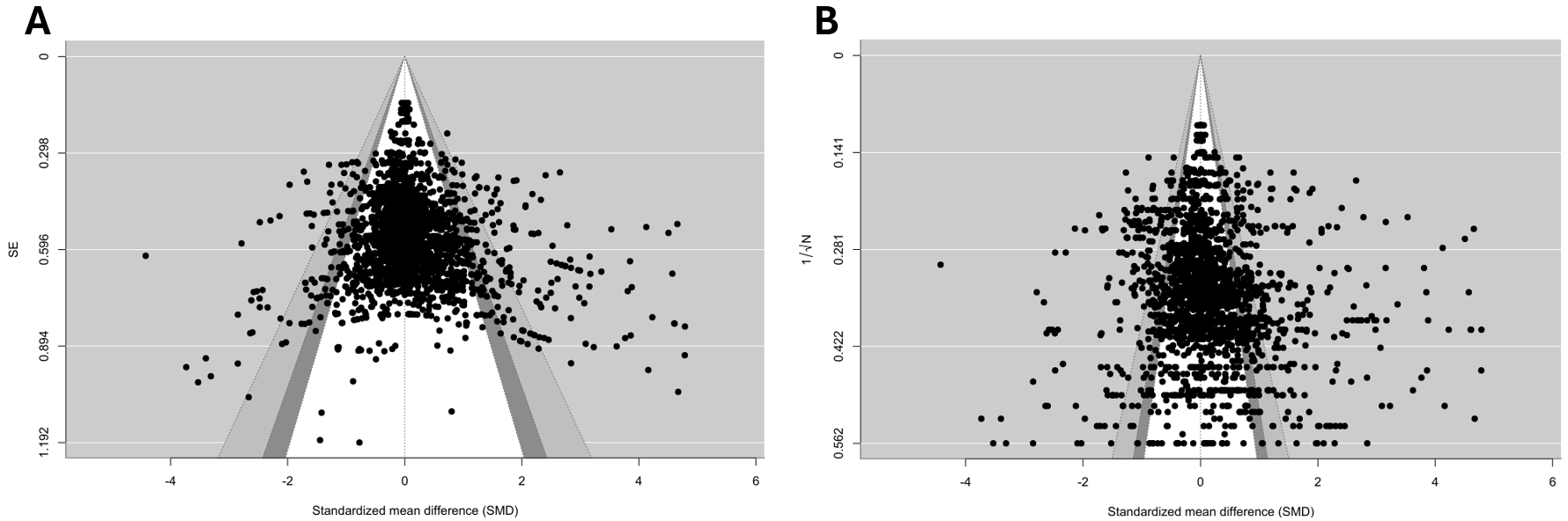

Supplementary Figure 10. Panel A demonstrates a traditional funnel plot of the effect measure against the standard error. Panel B demonstrates a funnel plot of the effect measure against a sample-size based measure of precision, as recommended for meta-analyses using the Standardised Mean Difference as the effect size measure (Zwetsloot et al. 2017).

#### Sensitivity Analysis: Results from a no-intercept model

The sensitivity analysis replicated the primary model in subgroups restricted to gene-drug pairings. Full model results are detailed in Supplementary Table 14, below. The results in the manuscript represent the difference between effect sizes for proximal and distal outcomes. Here, we report estimates for proximal and distal outcomes, separately for each of the subgroup analyses.

| **Subgroup** | **Term** | **Estimate** | **Standard Error** | ***p* value** |
| --- | --- | --- | --- | --- |
| CYP2D6 - Risperidone | Proximal | 0.5511 | 0.0747 | 2x10⁻⁸ |
| CYP2D6 - Risperidone | Distal | 0.3446 | 0.0570 | 2x10⁻⁵ |
| CYP2C19 - Escitalopram | Proximal | 0.5314 | 0.1269 | 0.006 |
| CYP2C19 - Escitalopram | Distal | 0.3184 | 0.1250 | 0.033 |
| CYP2D6 - Aripiprazole | Proximal | 0.6580 | 0.0848 | 2x10⁻⁵ |
| CYP2D6 - Aripiprazole | Distal | 0.3572 | 0.1532 | 0.055 |
| CYP2D6 - Haloperidol | Proximal | 0.5929 | 0.0641 | 1x10⁻⁵ |
| CYP2D6 - Haloperidol | Distal | 0.2991 | 0.1236 | 0.047 |

Supplementary Table 14. Results from sensitivity analyses (no-intercept models) testing whether pharmacogenomic effect sizes are moderated by outcome type across gene-drug pairings.

#### Secondary Analysis: Full Model Output

The secondary analysis expanded on the primary model by including an interaction term between outcome type (proximal vs distal) and metabolism phenotype (poor, intermediate, rapid, and ultra-rapid). An unstructured variance structure allowed between-study heterogeneity to vary across proximal and distal outcomes. Full model results are detailed in Supplementary Table 15, below.

| **Term** | **Estimate** | **Standard Error** | ***p* value** |
| --- | --- | --- | --- |
| Proximal * Poor Metaboliser | 1.0041 | 0.1054 | 1.36x10⁻¹² |
| Distal * Poor Metaboliser | 0.3807 | 0.0446 | 1.65x10⁻⁹ |
| Proximal * Intermediate Metaboliser | 0.4853 | 0.0402 | 6.71x10⁻²⁰ |
| Distal * Intermediate Metaboliser | 0.2913 | 0.0363 | 9.04x10⁻¹¹ |
| Proximal * Rapid Metaboliser | 0.1141 | 0.0891 | 0.235 |
| Distal * Rapid Metaboliser | 0.2623 | 0.0310 | 4.77x10^-5^ |
| Proximal * Ultrarapid Metaboliser | 0.3607 | 0.1040 | 0.0013 |
| Distal * Ultrarapid Metaboliser | 0.3373 | 0.0575 | 1.11x10^-5^ |

Supplementary Table 15. Full results from secondary analysis testing whether pharmacogenomic effect sizes are moderated by an interaction between outcome type and metabolism phenotype.

### PRISMA Checklist

| **Section and Topic** | **Item #** | **Checklist item** | **Location where item is reported** |
| --- | --- | --- | --- |
| **TITLE** | | |  |
| Title | 1 | Identify the report as a systematic review. | NA |
| **ABSTRACT** | | |  |
| Abstract | 2 | See the PRISMA 2020 for Abstracts checklist. | NA |
| **INTRODUCTION** | | |  |
| Rationale | 3 | Describe the rationale for the review in the context of existing knowledge. | Pg 1 – 2. |
| Objectives | 4 | Provide an explicit statement of the objective(s) or question(s) the review addresses. | Pg 1 – 2. |
| **METHODS** | | |  |
| Eligibility criteria | 5 | Specify the inclusion and exclusion criteria for the review and how studies were grouped for the syntheses. | Pg 4. |
| Information sources | 6 | Specify all databases, registers, websites, organisations, reference lists and other sources searched or consulted to identify studies. Specify the date when each source was last searched or consulted. | Pg 3. |
| Search strategy | 7 | Present the full search strategies for all databases, registers and websites, including any filters and limits used. | Supplementary materials Pg 2. |
| Selection process | 8 | Specify the methods used to decide whether a study met the inclusion criteria of the review, including how many reviewers screened each record and each report retrieved, whether they worked independently, and if applicable, details of automation tools used in the process. | Pg 4. |
| Data collection process | 9 | Specify the methods used to collect data from reports, including how many reviewers collected data from each report, whether they worked independently, any processes for obtaining or confirming data from study investigators, and if applicable, details of automation tools used in the process. | Pg 4 and 5. Supplementary materials Pg 3 – 5. |
| Data items | 10a | List and define all outcomes for which data were sought. Specify whether all results that were compatible with each outcome domain in each study were sought (e.g. for all measures, time points, analyses), and if not, the methods used to decide which results to collect. | Pg 5. |
|  | 10b | List and define all other variables for which data were sought (e.g. participant and intervention characteristics, funding sources). Describe any assumptions made about any missing or unclear information. | Pg 5. |
| Study risk of bias assessment | 11 | Specify the methods used to assess risk of bias in the included studies, including details of the tool(s) used, how many reviewers assessed each study and whether they worked independently, and if applicable, details of automation tools used in the process. | NA |
| Effect measures | 12 | Specify for each outcome the effect measure(s) (e.g. risk ratio, mean difference) used in the synthesis or presentation of results. | Pg 5. |
| Synthesis methods | 13a | Describe the processes used to decide which studies were eligible for each synthesis (e.g. tabulating the study intervention characteristics and comparing against the planned groups for each synthesis (item #5)). | NA |
|  | 13b | Describe any methods required to prepare the data for presentation or synthesis, such as handling of missing summary statistics, or data conversions. | Pg 4 and 5. Supplementary materials Pg 3 – 5. |
|  | 13c | Describe any methods used to tabulate or visually display results of individual studies and syntheses. | Pg 5. |
|  | 13d | Describe any methods used to synthesize results and provide a rationale for the choice(s). If meta-analysis was performed, describe the model(s), method(s) to identify the presence and extent of statistical heterogeneity, and software package(s) used. | Pg 5. |
|  | 13e | Describe any methods used to explore possible causes of heterogeneity among study results (e.g. subgroup analysis, meta-regression). | Pg 5. |
|  | 13f | Describe any sensitivity analyses conducted to assess robustness of the synthesized results. | Pg 5. |
| Reporting bias assessment | 14 | Describe any methods used to assess risk of bias due to missing results in a synthesis (arising from reporting biases). | Pg 5. |
| Certainty assessment | 15 | Describe any methods used to assess certainty (or confidence) in the body of evidence for an outcome. | NA |
| **RESULTS** | | |  |
| Study selection | 16a | Describe the results of the search and selection process, from the number of records identified in the search to the number of studies included in the review, ideally using a flow diagram. | Pg 4, Pg 6. |
|  | 16b | Cite studies that might appear to meet the inclusion criteria, but which were excluded, and explain why they were excluded. | NA |
| Study characteristics | 17 | Cite each included study and present its characteristics. | NA |
| Risk of bias in studies | 18 | Present assessments of risk of bias for each included study. | NA |
| Results of individual studies | 19 | For all outcomes, present, for each study: (a) summary statistics for each group (where appropriate) and (b) an effect estimate and its precision (e.g. confidence/credible interval), ideally using structured tables or plots. | NA |
| Results of syntheses | 20a | For each synthesis, briefly summarise the characteristics and risk of bias among contributing studies. | NA |
|  | 20b | Present results of all statistical syntheses conducted. If meta-analysis was done, present for each the summary estimate and its precision (e.g. confidence/credible interval) and measures of statistical heterogeneity. If comparing groups, describe the direction of the effect. | Pg 7 – 8. |
|  | 20c | Present results of all investigations of possible causes of heterogeneity among study results. | NA |
|  | 20d | Present results of all sensitivity analyses conducted to assess the robustness of the synthesized results. | Pg 8. |
| Reporting biases | 21 | Present assessments of risk of bias due to missing results (arising from reporting biases) for each synthesis assessed. | NA |
| Certainty of evidence | 22 | Present assessments of certainty (or confidence) in the body of evidence for each outcome assessed. | NA |
| **DISCUSSION** | | |  |
| Discussion | 23a | Provide a general interpretation of the results in the context of other evidence. | Pg 12 |
|  | 23b | Discuss any limitations of the evidence included in the review. | Pg 13 – 14. |
|  | 23c | Discuss any limitations of the review processes used. | NA |
|  | 23d | Discuss implications of the results for practice, policy, and future research. | Pg 12 – 14. |
| **OTHER INFORMATION** | | |  |
| Registration and protocol | 24a | Provide registration information for the review, including register name and registration number, or state that the review was not registered. | NA |
|  | 24b | Indicate where the review protocol can be accessed, or state that a protocol was not prepared. | NA |
|  | 24c | Describe and explain any amendments to information provided at registration or in the protocol. | NA |
| Support | 25 | Describe sources of financial or non-financial support for the review, and the role of the funders or sponsors in the review. | Pg 1. |
| Competing interests | 26 | Declare any competing interests of review authors. | Pg 1. |
| Availability of data, code and other materials | 27 | Report which of the following are publicly available and where they can be found: template data collection forms; data extracted from included studies; data used for all analyses; analytic code; any other materials used in the review. |  |

*From:* Page MJ, McKenzie JE, Bossuyt PM, Boutron I, Hoffmann TC, Mulrow CD, et al. The PRISMA 2020 statement: an updated guideline for reporting systematic reviews. BMJ 2021;372:n71. doi: 10.1136/bmj.n71. This work is licensed under CC BY 4.0. To view a copy of this license, visit <https://creativecommons.org/licenses/by/4.0/>
